# Post-pandemic changes in population immunity have reduced the likelihood of emergence of zoonotic coronaviruses

**DOI:** 10.1101/2025.03.17.25323820

**Authors:** Ryan M Imrie, Laura A Bissett, Savitha Raveendran, Maria Manali, Julien A R Amat, Laura Mojsiejczuk, Nicola Logan, Andrew Park, Marc Baguelin, Mafalda Viana, Brian J Willett, Pablo R Murcia

## Abstract

Infections caused by endemic viruses, and the vaccines used to control them, often provide cross-protection against related viruses. This cross-protection has the potential to alter the transmission dynamics and likelihood of emergence of novel zoonotic viruses with pandemic potential. Here, we investigate how changes in population immunity after the COVID-19 pandemic have impacted the likelihood of emergence of a novel sarbecovirus, termed SARS-CoV-X. We show that sera from patients with different COVID-19 immunological histories possess cross-neutralising antibodies against the spike (S) protein of multiple zoonotic sarbecoviruses. Mathematical simulations using these viruses show a significant reduction in their likelihood of emergence in populations with current levels of SARS-CoV-2 natural and vaccine-derived immunity, with the outcome determined by the extent of cross-protection and the *R*_0_ of the novel virus. We also show that preventative vaccination programs against SARS-CoV-X using currently available COVID-19 vaccines can help resist emergence even in the presence of co-circulating SARS-CoV-2. However, it was possible for a theoretical vaccine with very high specificity to SARS-CoV-2 (i.e., one which elicits very low cross-protection against SARS-CoV-X) to increase the likelihood of SARS-CoV-X emergence, due to its effects on SARS-CoV-2 prevalence and, by extension, the levels of naturally-derived cross-protection in the human population. Overall, our findings show that SARS-CoV-2 circulation and COVID-19 vaccination have generated widespread population immunity against antigenically related sarbecoviruses, and this new immunological barrier is likely to be a strong determinant of the ability of novel sarbecoviruses to emerge in humans.

**One-Sentence Summary:** Future emergence of zoonotic coronaviruses is less likely due to post-pandemic shifts in population immunity.

## Introduction

Identifying viruses with zoonotic potential before they emerge is essential for pandemic preparedness. To infect humans, zoonotic viruses must overcome multiple biological barriers. As intracellular pathogens, viruses rely on host machinery for replication, but molecular incompatibilities between viral and host proteins can restrict infection. Humans also have antiviral defenses, including host restriction factors that inhibit viral replication [1], immune cells such as natural killer (NK) cells and cytotoxic T cells (CTLs) that eliminate infected cells [2,3], and interferons that suppress viral replication while promoting immune responses [4]. Additionally, neutralizing antibodies, induced by infection or vaccination, limit viral replication and aid recovery from infection [5]. While overcoming these barriers is required for successful zoonoses, not all zoonotic viruses pose a pandemic risk. This is because sustained human-to-human transmission is a prerequisite for viral emergence, and thus the risk of emergence by viruses that are either non-transmissible or have limited transmissibility is considered low [6].

Severe acute respiratory syndrome coronavirus (SARS-CoV), Middle East respiratory syndrome coronavirus (MERS-CoV), and severe acute respiratory syndrome coronavirus 2 (SARS-CoV-2) are highly pathogenic coronaviruses that have transmitted from animal hosts to humans in the 21^st^ century [7]. While subsequent human-to-human transmission has been reported for all of them, their epidemiological trajectories differed. SARS-CoV emerged in China in 2002 and caused a large international outbreak across 27 countries, with 8096 reported cases and 774 deaths [8]. By 2003 the SARS epidemic was controlled and only five additional SARS-CoV zoonotic infections have been reported since then [9]. Most cases of MERS-CoV infection are the result of individual zoonotic events with limited onward transmission [10]. As of February 2025, there have been 2626 confirmed cases of MERS [11], and in the largest outbreak 186 cases were diagnosed [12]. In contrast, SARS-CoV-2 emerged in humans in December 2019, resulting in a global pandemic with more than 770 million confirmed cases and over 7 million deaths as of February 2025. Within a year since the COVID-19 pandemic was declared, SARS-CoV-2 vaccines were developed, and vaccination programs were implemented globally [13]. As of August 2024, over 13 billion vaccine doses have been administered worldwide [14].

Mutations in the spike (S) protein of SARS-CoV-2 have enabled the virus to escape immunity induced by vaccination or infection, resulting in epidemic waves where reinfections and vaccine breakdowns are common [15]. SARS-CoV-2 continues to circulate even in populations with high vaccination coverage [16], although the number of hospitalizations and deaths has decreased significantly compared to the pre-vaccine era [17]. As a result, the global immunological landscape has changed dramatically since the emergence of COVID-19, as a large proportion of the human population has developed anti-SARS-CoV-2 antibodies. For example, in Scotland alone, it is estimated that the proportion of SARS-CoV-2 seropositive individuals increased from 4.4% in May 2020 to 96.5% in June 2022 [18].

Cross-immunity can impact the transmission dynamics of genetically related respiratory viruses, as illustrated by the dynamic patterns of human respiratory syncytial virus, metapneumovirus, parainfluenza viruses and enteroviruses [19,20]. While recent reports explored the effectiveness of SARS-CoV-2 immunity against infection, hospitalization, or death by different SARS-CoV-2 variants [21,22], no studies have investigated the impact of SARS-CoV-2 cross-immunity on the likelihood of emergence of novel zoonotic viruses. Various surveillance and virus discovery studies have shown that viruses belonging to the same subgenus as SARS-CoV and SARS-CoV-2 (sarbecoviruses) circulate in wildlife over broad geographic regions [23,24]. Examples include the bat viruses Rs4084, RaTG13, BANAL-52, BANAL-103, BANAL-114, BANAL-236, BANAL-247 (all the BANAL CoVs can bind the human ACE2 receptor [25]), as well as the pangolin viruses GD-1, GX-P5L and GX/P1E [26].

A key priority for pandemic preparedness is to develop methodologies to assess the pandemic risk of animal-origin viruses [27] including any potential zoonotic sarbecoviruses, which we refer to collectively as “SARS-CoV-X”. We hypothesized that post-pandemic changes in population immunity have reduced the likelihood of antigenically related zoonotic sarbecoviruses emerging in humans. To test this hypothesis, we first measured the extent to which SARS-CoV-2 antibodies cross-neutralised a panel of related sarbecoviruses using sera from four groups: individuals with no history of SARS-CoV-2 infection or vaccination, those who had recovered from infection, those who had been vaccinated, and those with immunity from both infection and vaccination (i.e., hybrid immunity). The sarbecovirus panel included two close relatives of SARS-CoV-2 – the bat virus RaTG13 and the pangolin virus GX/P1E – as well as SARS-CoV and its close relative, the bat virus RS4084. To assess how cross-immunity influences the likelihood of SARS-CoV-X emergence, we constructed an age-stratified stochastic SEIRS (Susceptible - Exposed - Infectious - Recovered – Susceptible) model, parameterized to reflect the population of Scotland. This model simulated the co-circulation of SARS-CoV-2 and a hypothetical SARS-CoV-X under varying conditions of vaccination uptake, cross-immunity, and different SARS-CoV-X basic reproduction numbers (*R*_0_). By modeling these scenarios, we quantified the extent to which existing immunity and vaccination mitigate the pandemic potential of zoonotic sarbecoviruses. Additionally, as virus surveillance efforts aim to act as an early-warning system for potential emergence events [28], and widespread vaccination with a cross-reactive vaccine is a possible preventative action [29], we explored how a 2-month preventative vaccination program using available COVID-19 vaccines could impact the likelihood of sarbecovirus emergence in the presence of SARS-CoV-2 co-circulation.

## Results

### SARS-CoV-2 infection and vaccination produce cross-reactive antibodies against other zoonotic sarbecoviruses

The neutralising activity in sera from individuals with different histories of SARS-CoV-2 natural infection and vaccination was quantified *in vitro* using lentiviral pseudotypes carrying a luciferase reporter gene and the spike proteins of four sarbecoviruses: SARS-CoV, Rs4084 (bat), GX/P1E (pangolin), and RaTG13 (bat) (Figure 1A-C). Sera from naïve individuals showed the lowest levels of neutralisation across the four viruses (mean = 14.8%, SE = 11.2). Neutralising activity increased significantly in sera from individuals with known SARS-CoV-2 natural infection (mean = 41.7%, SE = 10.8, p < 0.01); those who had received 1 vaccine dose (mean = 45.2%, SE = 11.3, p < 0.01); and those who had received two vaccine doses (mean = 54.5%, SE = 11.2, p < 0.01). Neutralising activity increased further in sera from individuals with hybrid immunity, who had both recovered from natural infection and received one dose (mean = 64.1%, SE = 10.9, p <0.01) or two doses of vaccine (mean = 67.0%, SE = 10.8, p <0.01). The extent of cross-neutralisation also varied between viruses, with the lowest value across immune groups seen for SARS-CoV (μ =29.9%, SE = 13.2). Significant increases in cross-neutralisation were observed for Rs4084 (μ = 39.7% ± 13.2, p < 0.01), GX/P1E (μ = 43.2%, SE = 13.2, p < 0.01), and RaTG13 (μ = 78.6%, SE = 13.2, p < 0.01) (Supplementary Tables 3-4). These values are consistent with a decrease in cross-neutralisation with increasing evolutionary distance from the SARS-CoV-2 (Wuhan-Hu-1) spike protein (Figure 1D). Accordingly, a significant positive relationship between spike protein amino acid similarity (%) and cross-neutralisation was detected in these data (*β*^2^= 0.09, SE = 0.003, p < 0.01) (Figure 1E, Supplementary Tables 5-6). These results indicate that antibodies elicited against SARS-CoV-2 can cross-neutralise other sarbecoviruses, and that the level of cross-neutralisation is influenced by the route of immune acquisition (natural infection, vaccination, or hybrid) and the similarity of the spike proteins of the immunising and infecting viruses.

**Figure 1:**
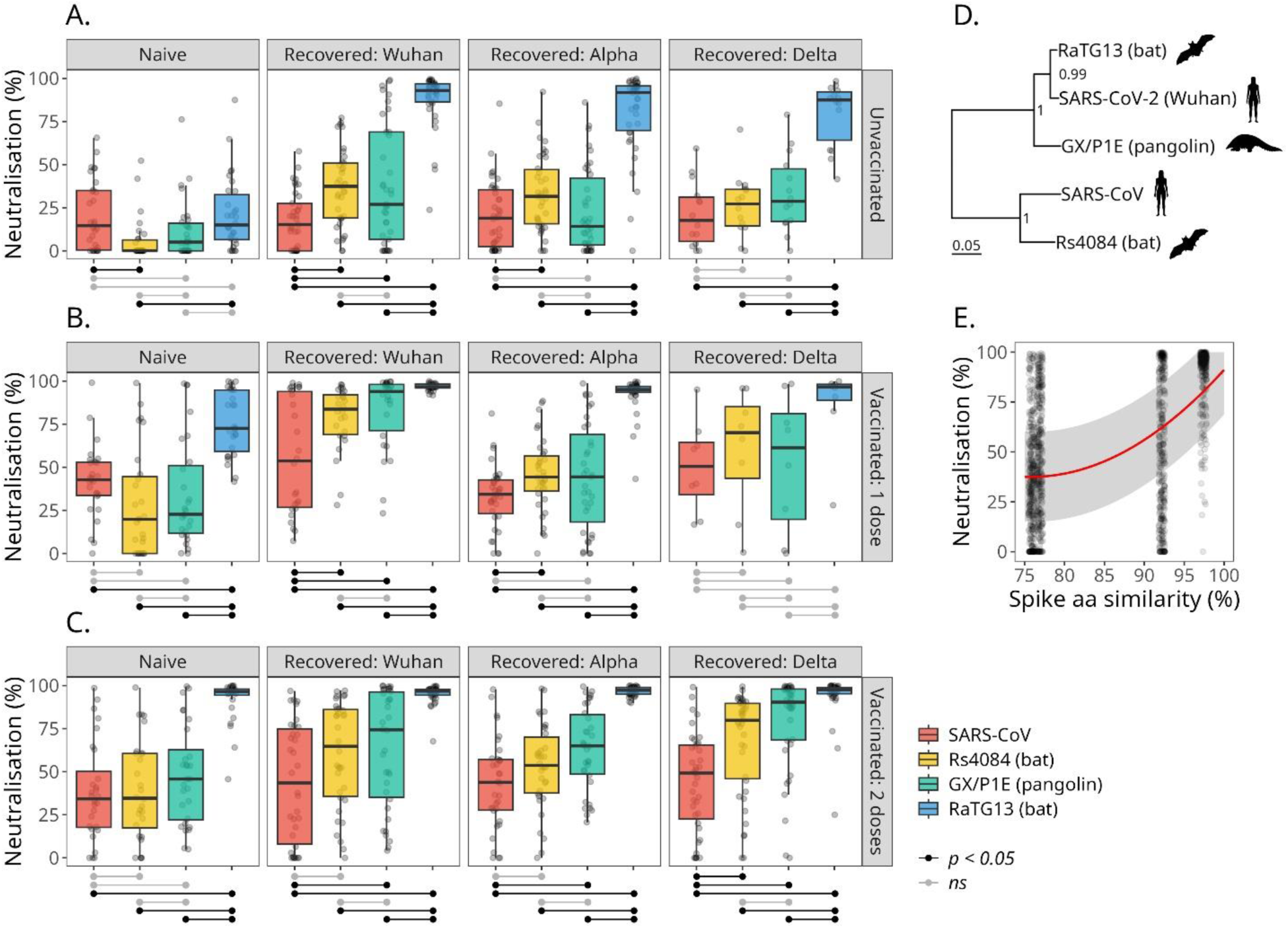
Neutralisation of viral pseudotypes carrying the spike proteins of different sarbecoviruses by sera from individuals of different infection and vaccination histories. Boxplots show the percentage neutralisation of viral pseudotypes by sera from individuals who were unvaccinated (A), vaccinated once (B), or vaccinated twice (C) against SARS-CoV-2. Results are separated into subplots (columns) based on an individual’s history of natural infection with SARS-CoV-2, and separate boxplots are shown for neutralisation of viral pseudotypes carrying the spike protein of SARS-CoV (red), Rs4084 (yellow), GX/P1E (green) or RaTG13 (blue). Significant differences in neutralising activity are shown with black horizontal lines below each subplot, assessed using Welch’s t-tests with Holm correction for multiple testing. D) A maximum clade credibility phylogeny inferred from the spike protein sequences of each SARS coronavirus and SARS-CoV-2 (Wuhan-Hu-1), with posterior probabilities of each clade and a scale bar demonstrating amino acid substitutions per site per unit time. E) The relationship between percentage neutralisation and spike protein sequence similarity (%) to SARS-CoV-2 (Wuhan-Hu-1) modelled as a quadratic function. Data points are neutralisation data taken from A-C (excluding Naive + Unvaccinated individuals). The trend line and shaded area indicate the predicted values and standard error, respectively, from a quadratic mixed-effects model, with random effects of vaccine history, natural infection history, and serum ID.

### Population immunity against SARS-CoV-2 decreases the likelihood of emergence of zoonotic sarbecoviruses

To investigate the likelihood of zoonotic sarbecovirus emergence under varying conditions of cross-protection and vaccination, as well as different values of *R*_0_, we developed an age-stratified stochastic SEIRS model (Supplementary Figure 1) parameterised to resemble the projected demographic structure of Scotland from 2020-2028. The model included separate EIR compartments for SARS-CoV-2 and SARS-CoV-X infection for each immune group (each combination of vaccination status and infection history) in the pseudotype neutralization data (Figure 1). Each group was given unique transmission probabilities, scaled by the complement (1 − *x*) of cross-neutralisation, making individuals with higher immune cross-neutralisation less likely to become infected when contacting an infectious person. Vaccination rates from the initial COVID-19 vaccination program and spring/winter booster programs, along with waning immunity calculated from the half-lives of vaccine-derived antibodies, produced a dynamic immune landscape of vaccine protection and cross-protection over time, peaking in winter at ∼12–19% coverage and dominated by individuals aged 65+ (Supplementary Figure 2). The model accurately tracked real-world SARS-CoV-2 prevalence until the initial Omicron wave (Figure 2A), after which the availability of detailed serological, vaccination, and social contact data decreased as the UK moved into the “post-COVID” era. After this point, the model stabilized at a SARS-CoV-2 prevalence of 2.2%, consistent with current estimates, but did not capture short-term fluctuations seen in real-world data.

**Figure 2.**
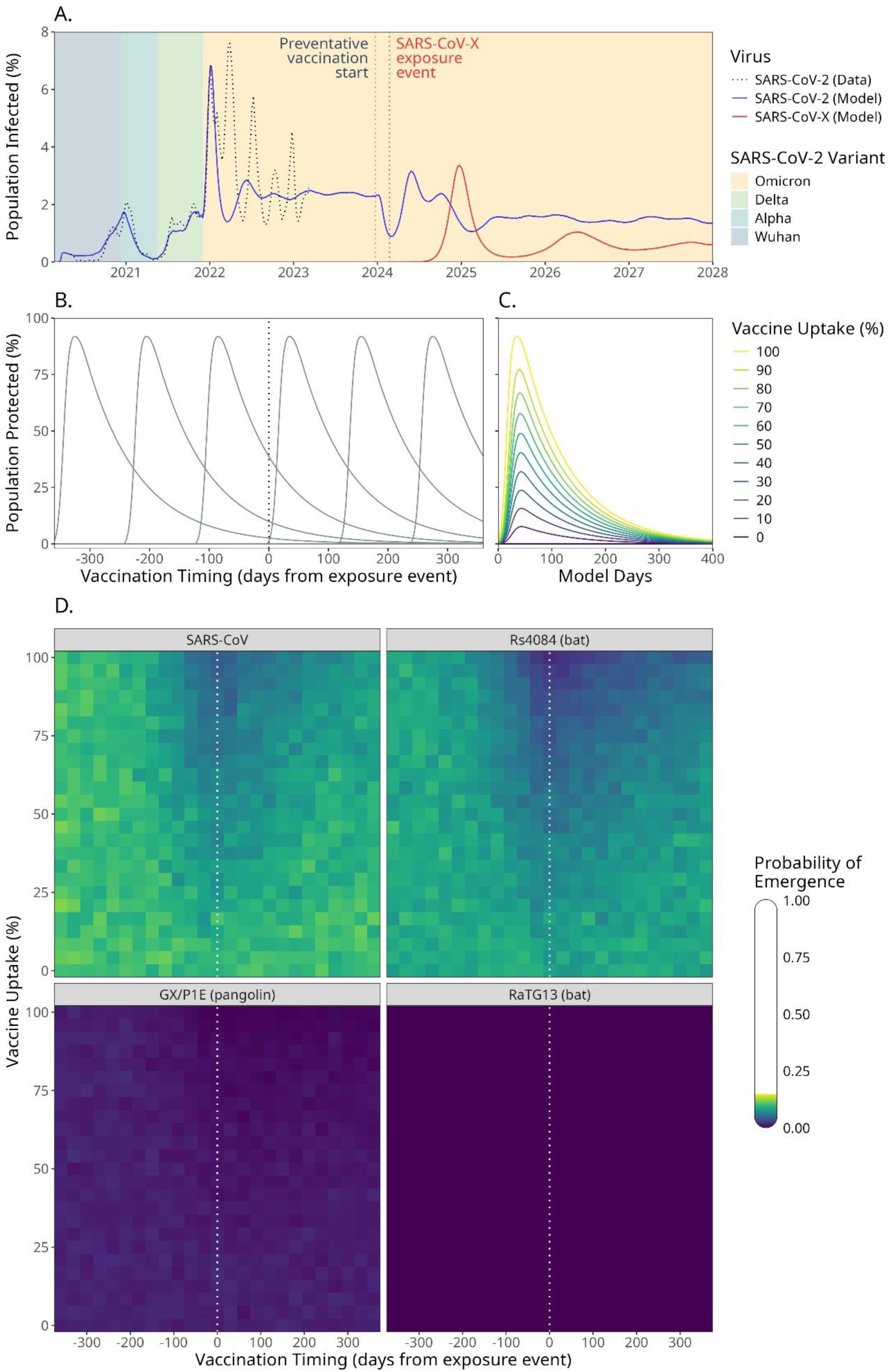
(overleaf): Probability of emergence of different SARS coronaviruses in the presence of vaccination and co-circulating SARS-CoV-2. A) An example model run showing percentages of individuals infected with SARS-CoV-2 (blue) and SARS-CoV-X (red) over time. The predominant SARS-CoV-2 variant is indicated by the shaded background area, and estimates of real SARS-CoV-2 prevalence, taken from the UK Coronavirus Infection Survey, are shown as a dotted black line. In this example, preventative vaccination began 30 days before SARS-CoV-X emergence, with a vaccine uptake of 100%, and SARS-CoV-X phenotypes were set to those of SARS-CoV. A model run where SARS-CoV-X emerged successfully is shown for illustrative purposes, although this may not represent the most likely outcome given these model conditions. The percentage of the population protected by preventative vaccination depends on the timing (B) and uptake (C) of the vaccination campaign. D) Heatmaps display the predicted probability of emergence for four SARS coronaviruses in a population with co-circulating SARS-CoV-2 under varying conditions of preventative vaccination.

The model was used to investigate the likelihood of a novel sarbecovirus emerging in a human population with a) current levels of SARS-CoV-2 circulation and COVID-19 vaccination, and b) during a theoretical 2-month preventative vaccination program, where currently available COVID-19 vaccines were used to resist SARS-CoV-X emergence (Figure 2B-C). As no *R*_0_ estimates are available for the zoonotic sarbecoviruses in a comparable human population to the model, this parameter was set to the model-fitted estimate for SARS-CoV-2 (Wuhan), which had an *R*_0_of 1.57. Our simulations showed that all sarbecoviruses included in the neutralisation experiments displayed a low probability of emergence under current conditions (Figure 2D). The highest probability was observed for SARS-CoV (11.1%), followed by Rs4084 (9.9%), GX/P1E (2.0%), and RatG13, which failed to emerge once in 50,000 model runs. Preventative vaccination had no detectable effect on the probability of emergence when vaccination started >200 days before the date of the first SARS-CoV-X case. However, a pronounced reduction on the emergence probability of SARS-CoV and Rs4084 was observed across all other timings (180 days before to 360 days after the date of the first SARS-CoV-X human case), and a more modest effect was seen in GX/P1E as vaccination pushed its probability of emergence close to zero (Supplementary Figure 3). The most effective vaccine programs began at the point of SARS-CoV-X introduction and reduced the probability of emergence from 11.6% to 4.3% for SARS-CoV; 8.4% to 3.1% for Rs4084; and from 0.8% to 0.3% for GX/P1E. These results show that short-term and widespread vaccination with currently available SARS-CoV-2 vaccines can be used to reduce the emergence potential of novel sarbecoviruses even in the presence of co-circulating SARS-CoV-2.

To further characterise the effects of cross-protection and preventative vaccination on the probability of SARS-CoV-X emergence, model runs were performed for nine hypothetical sarbecoviruses with variable *R*_0_ values (2, 3, and 4) under conditions where cross-protection provided by SARS-CoV-2 infection and vaccination were equal and set at 33%, 67%, or 100% (Figure 3, Supplementary Figure 4). In the absence of preventative vaccination the probability of emergence increased with *R*_0_ and decreased with higher levels of cross-immunity, such that the probability of emergence of a virus with cross-reactivity of 33% and an *R*_0_ of 2 (34.0%) was similar to that of a virus with cross-reactivity of 67% and an *R*_0_ of 3 (39.8%), and that of a virus with cross-reactivity of 100% and an *R*_0_of 4 (35.8%). Preventative vaccination was most effective at high levels of vaccine cross-protection and against viruses of lower *R*_0_, and these effects were more evident in model explorations where vaccine effectiveness varied independently from cross-immunity to SARS-CoV-2 (Supplementary Figures 5-6). Here, the emergence of a virus with vaccine cross-reactivity of 100% and an *R*_0_ of 2 was reduced in vaccination programs begun 150 days before to 60 days after the date of the first SARS-CoV-X human case, and at their optima these programs reduced the probability of emergence from 28.1% to 4.3%. In contrast, the probability of emergence of a virus with an *R*_0_= 4 was only reduced in programs begun 120 days before the first human infection and had no detectable impact when started after the first SARS-CoV-X case, although well-timed vaccination was still able to reduce the probability of emergence of this virus from 67.8% to 35.1%.

**Figure 3:**
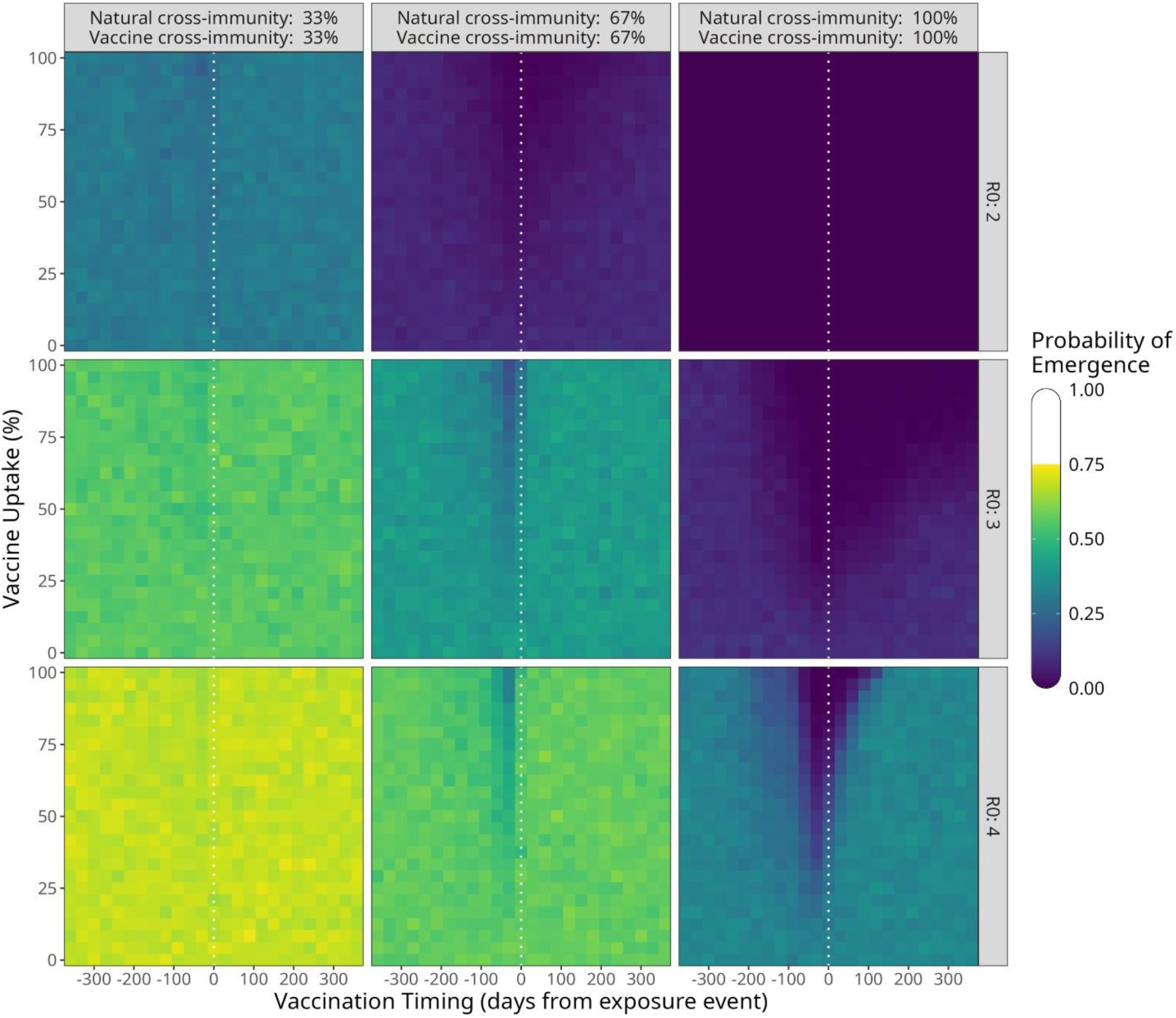
Probability of emergence of theoretical SARS coronaviruses under conditions of equal natural and vaccine cross-immunity. Heatmaps show point estimates of the probability of emergence for nine theoretical SARS coronaviruses with different *R*_0_values (panel rows) and varying conditions of cross-immunity and vaccine effectiveness (panel columns) in a population with co-circulating SARS-CoV-2. In these scenarios, protection against SARS-CoV-X infection conferred from recovering from natural infection with SARS-CoV-2 (“natural cross-immunity”) and vaccination (“vaccine cross-immunity”) are identical.

### A theoretical vaccine with high SARS-CoV-2 specificity can increase the likelihood of SARS-CoV-X emergence

As cross-immunity from both vaccination and natural infection with SARS-CoV-2 can decrease the likelihood of SARS-CoV-X emergence, it may be possible for a highly specific SARS-CoV-2 vaccine with little cross-reactivity to *increase* the probability of SARS-CoV-X emergence, provided it reduces SARS-CoV-2 prevalence enough to lower the level of natural cross-immunity in the population. This outcome was visible in our model when vaccine cross-reactivity was set very low (5%) while cross-reactivity elicited by natural infection with SARS-CoV-2 was high (Figure 4). The detrimental effect of vaccination under these conditions became more pronounced as *R*_0_decreased and natural cross-immunity increased (Supplementary Figure 7). For example, under conditions of 67% natural cross-immunity, the probability of emergence increased with vaccination by up to 7.5% for a virus of *R*_0_= 4, 10.2% for a virus of *R*_0_= 3, and 14.1% for a virus of *R*_0_ = 2. The strongest detected negative effect of vaccination was under conditions of 100% natural cross-immunity and a virus of *R*_0_= 3, where the probability of SARS-CoV-X emergence increased by 21.8%. Even in the presence of detrimental vaccination, a virus with an *R*_0_ of 2 remained incapable of emerging under conditions of 100% natural cross-immunity to SARS-CoV-2. Notably, in an additional run under conditions of 79% natural cross-immunity, vaccination with a 5% cross-reactive vaccine allowed a virus with no detectable ability to emerge in the human population to emerge in 3.2% of trials (Supplementary Figure 8).

**Figure 4:**
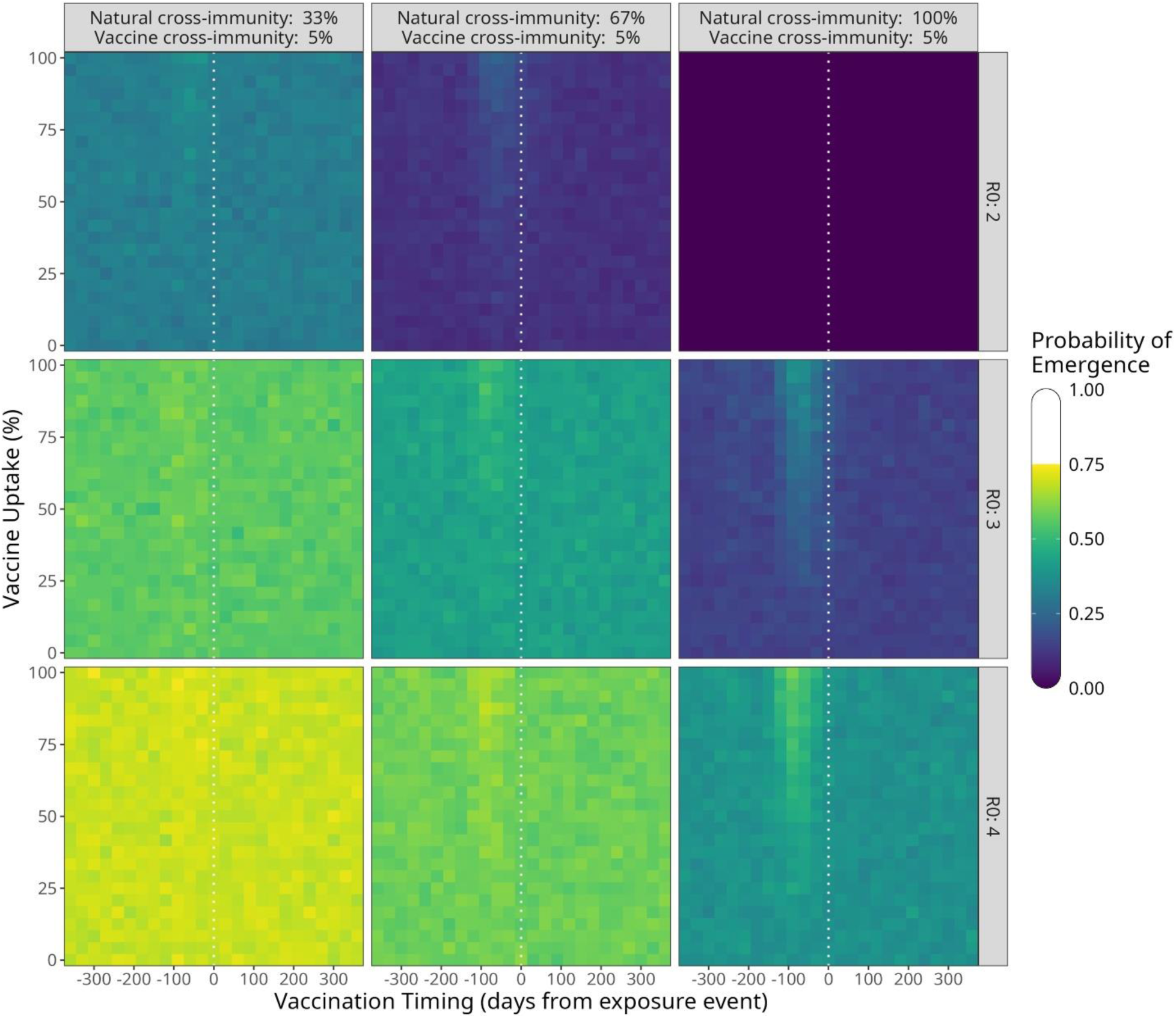
Probability of emergence of theoretical SARS coronaviruses under conditions of low vaccine cross-immunity and high natural cross-immunity. Heatmaps show point estimates of the probability of emergence for three theoretical SARS coronaviruses with different *R*_0_values (panel rows) and varying conditions of natural cross-immunity (panel columns) in a population with co-circulating SARS-CoV-2, under and low (5%) vaccine cross-immunity.

## Discussion

The emergence of COVID-19 illustrates the potentially devastating effects of viral zoonoses, and preparations for future viral emergence events will require effective approaches to quantify the risk of emergence and the impact of intervention strategies. Here, we have investigated the likelihood of a new zoonotic sarbecovirus emerging in the post-pandemic era with current levels of natural cross-immunity from SARS-CoV-2 circulation and vaccine-derived immunity from COVID-19 vaccination. We show that the emergence of new sarbecoviruses is likely to be resisted considerably by cross-reactivity of the adaptive immune response targeting SARS-CoV-2, and that close relatives of SARS-CoV-2 such as RaTG13 may be currently incapable of emerging in the human population. These results are consistent with modelling studies showing that cross-immunity elicited by human influenza viruses can lower the attack rate of avian-origin influenza viruses with pandemic potential [30]. Using already available COVID-19 vaccines as a preventative measure – both pre-emptively and as a reaction to SARS-CoV-X exposure – led to a further reduction in the likelihood of a novel sarbecovirus becoming endemic in the human population, with programs targeting viruses with similar *R*_0_ to SARS-CoV-2 Wuhan (1.57) showing beneficial effects up to 1 year after the first SARS-CoV-X case.

To measure the breadth and potency of the cross-protective antibody response, we performed pseudotype-based neutralisation assays using virions carrying the spike proteins of four sarbecoviruses closely related to SARS-CoV-2, including SARS-CoV, a known zoonotic and transmissible virus. These analyses are particularly relevant given that neutralising antibody titers have been proposed as correlates of protection against different viral diseases [31], including COVID-19 [32], measles [33], and smallpox [34]. Our data suggest that the level of cross-neutralisation between the spike proteins of SARS-CoV-2 and zoonotic sarbecoviruses is associated with their phylogenetic distance. However, this seems at odds with the low neutralisation levels observed against Omicron BA.1.17, despite the latter sharing 96.9% spike amino acid identity with the SARS-CoV-2 Wuhan strain (Supplementary Figure 9). This disparity has also been observed by others [35] and attributed to differences in immune selection pressures as SARS-CoV-2 variants, but not animal sarbecoviruses, evolve to escape prior immunity. This discrepancy is also explained by antigenic cartography studies showing that a small number of mutations in the receptor binding domain of the spike protein can have a large antigenic effect [36].

The highest levels of cross-neutralisation across the four sarbecoviruses tested were consistently observed in patients with hybrid immunity (i.e. those infected and vaccinated), suggesting that vaccine-breakdown infections by immune evasive SARS-CoV-2 variants may, ultimately, have a strong protective effect against subsequent SARS-CoV-X infection, and that vaccination should be encouraged even in patients with a history of prior infection. In unvaccinated individuals with immunity acquired by natural infection alone, the breadth and potency of antibody-mediated neutralisation was lower than in patients with hybrid immunity and was determined by the SARS-CoV-2 infecting strain. This is consistent with findings that protection conferred by natural infection varies over time and is influenced by the antigenic evolution of SARS-CoV-2, with pre-Omicron infections offering durable immunity, whereas immunity following Omicron infection wanes more rapidly, likely due to increased immune escape [37]. As a result, levels of cross-protection in individuals who are unvaccinated – or whose vaccine-derived antibodies have waned – might fluctuate according to the antigenic evolution and epidemiology of SARS-CoV-2. Accordingly, future genomic and serological surveillance should continue to quantify the extent of cross-neutralisation elicited by new SARS-CoV-2 variants as they arise.

It is conceivable that a zoonotic sarbecovirus could increase its prevalence in animal populations (like H5N1 influenza). Our results show that preemptive vaccination with available COVID-19 vaccines in high-risk areas could be used to control SARS-CoV-X emergence before any human cases arise. Unlike the variable immunity conferred by natural infection with successive variants of SARS-CoV-2, immunity elicited by vaccination should be more uniform. Hence, the breadth of cross-protection obtained by vaccination is likely to be more consistent and predictable. The first generation of COVID-19 vaccines were derived predominantly from the ancestral Wuhan virus, but recent formulations of bivalent vaccines include both the Wuhan and Omicron strains. It is possible that such bivalent vaccines will elicit a broader cross-neutralising response. While recent studies have indicated that immunological imprinting may constrain the ability of bivalent vaccines to broaden the immune response [38,39], others have shown that vaccines designed to target conserved regions of the sarbecovirus spike can elicit broader and more cross-reactive immune responses, improving protection against SARS-CoV-2 variants and related viruses [40,41]. Another important factor to consider is the type of vaccine used. For example, it has been shown that mRNA-based vaccines elicit higher neutralisation titres than adenovirus-vectored vaccines (i.e., ChAdOX vaccine), and in some cases higher titres than natural infections [42]. These findings highlight the potential benefits to preparedness and prevention in developing cross-protective, universal vaccines such as those proposed for pan-sarbecoviruses [43], but also of other antigenically variable pathogens such as influenza viruses [44].

Our study has various limitations. In the absence of more detailed information, we assumed that the *in vitro* activity of neutralizing antibodies correlates linearly to *in vivo* protection from infection, although this direct proportionality may not be true in all cases [5], and thus we also included theoretical explorations where cross-immunity and vaccine effectiveness is unlinked from the neutralization assay data. Our model itself is conceptual, and more work will be needed to perform accurate risk assessments of emergence potential. For example, future models should include the contribution of cross-immunity by cellular responses such as T cells [45]; spatial and demographic variation in social behaviour and vaccine uptake and compliance [46]; and more realistic changes in the immunological landscape over time as vaccine technologies change and new virus variants emerge [47]. Increases in the complexity of the modelling approach should be matched by improvements in the techniques used to quantify virus infection phenotypes and predict the likely trajectories of virus evolution, as any actionable risk assessment of the likelihood of virus emergence will rely on the combined insights of each of these elements.

Identifying pathogens that might cause future pandemics is essential for preparedness [27]. The current H5N1 influenza panzootic [48] and the increasing number of human infections by zoonotic H5N1 [49] highlight the importance and timeliness of this issue, as does the continuous identification of novel animal coronavirus able to effectively bind human ACE2 receptors [50]. Since 2017, the World Health Organization (WHO) has published a list of priority pathogens and diseases with the potential to cause future pandemics, including SARS-CoV. Together, our results indicate that the risk of emergence of novel sarbecoviruses antigenically related to SARS-CoV-2 has decreased due to shifts in population immunity in the post-pandemic era, and that endemic viruses in humans may act as a strong barrier to the emergence of novel zoonotic viruses.

## Materials and Methods

### Serum samples

Residual biochemistry serum samples were randomly collected from both primary care (general practice) and secondary care (hospital) settings by the NHSGGC Biorepository between March 31, 2020, and September 22, 2021. Metadata including date of collection, sex, age, and vaccination history were provided, and the natural infection history of each patient was inferred from the date of last positive PCR test. Of the approximately 41,000 samples collected, a representative subsample of 350 sera were selected that maintained the underlying demographic proportions of the full sample.

### Generation of lentiviral pseudotypes

Gene constructs containing the SARS-CoV (GenBank: AY394995.1), Rs4084 (GenBank: KY417144.1), GX/P1E (GISAID: EPI_ISL_410539), and RaTG13 (GenBank: MN996532.2) spike genes were codon-optimized and synthesized by GenScript Biotech. Each construct was cloned into an expression vector and co-transfected with p8.91-HIV (a gag-pol lentiviral core [51]) and pCSFLW-firefly (which contains a luciferase reporter gene with HIV-1 packaging signal [52]) into HEK293T cells using polyethylenimine. Cells were maintained at 37°C, 5% CO_2_, in Dulbecco’s modified Eagle’s medium (DMEM) supplemented with 10% fetal bovine serum (FBS), 2-mmol/l L-glutamine, 100µg/ml streptomycin and 100IU/ml penicillin. Supernatants containing HIV (SARS-CoV-X) virions were harvested 48 hours after transfection and frozen at -80°C before use.

### Neutralization Assays

Pseudotype based neutralization assays were performed as described previously [53,54]. Briefly, stably expressing HEK293-hACE2 (human angiotensin-converting enzyme 2) cells were generated by transfection of HEK293 cells with pSCRPSY-hACE2 and maintained as above. Serum samples were diluted 1:25 in Complete DMEM, and 25µl of each diluted serum sample was transferred to the wells of a white 96-well plate. An equal volume (25µl) of viral pseudotype was added to each well, resulting in a final serum concentration of 1:50. Plates were incubated for 1 hour at 37°C, 5% CO_2_, after which 50µl of 4×10^5^ cells/ml of HEK293-hACE2 cells were added to each well. Plates were then incubated for a further 48 hours. After incubation, 100µl of Steadylite Plus was added to each well, and the plate was incubated in the dark for 10 minutes. Luciferase activity in each well was quantified on a Perkin Elmer Ensight multimode plate reader. The proportion of pseudovirus neutralization was calculated as 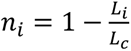, where *L*_*i*_ is the luciferase activity of sample wells, and *L*_*c*_ the luciferase activity of no-serum control wells. Two technical replicates for each combination of serum and pseudovirus were collected and averaged to provide final measures of neutralization. Where *n*_*i*_ < 0, which occurred when the luciferase activity of sample wells exceeded the activity of no-serum control wells, the proportion of neutralization was set to zero.

To estimate overall effects of vaccination and recovery from SARS-CoV-2 infection on neutralization, data from all serum groups were used to fit linear mixed effects models with a fixed effects structure *y*_*i*,*r*,*v*_ = *β*_1,*r*_ + *β*_2,*v*_ + *β*_3,*r*,*v*_ + *e*. Here, *y*_*i*,*r*,*v*_ is the percentage neutralization of serum from individual *i* with recovery status *r* and vaccine status *v*. *β*_1,*r*_, *β*_2,*v*_, and *β*_3,*r*,*v*_ represent the predicted contribution to percentage neutralization of recovery status, vaccination status, and their interaction, respectively, while *e* represents the model residuals. In this model, random effects of pseudotype virus identity and serum ID were included to account for between-group variability and repeated measures from individual sera. Differences in neutralization between viruses were estimated using a similar modelling approach but with virus identity included as a fixed effect and recovery and vaccine status as random effects. To estimate the overall effect of spike protein similarity on neutralization, data from all serum groups, excluding naïve + unvaccinated individuals, were used to fit a quadratic mixed effects model with a fixed effects structure *y*_*i*,*s*_ = *β*_*s*_^2^ + *e*. In this model, *y*_*i*,*s*_ is the percentage neutralization of serum from individual *i* against spike protein of similarity *s*; *β*_*s*_^2^ represents the predicted percentage neutralization against a spike protein of similarity *s*. Random effects were included for recovery status, vaccination status, and serum ID. Similarity were calculated as the percentage amino acid identity to the Wuhan-Hu-1 spike protein sequence, taken from a translation alignment (described below).

### Spike protein phylogeny

Spike protein coding sequences of SARS-CoV-2 (Wuhan-Hu-1 strain, GenBank: MN908947.3), SARS-CoV, Rs4084, GX/P1E, and RaTG13 (accessions in the previous section) were translated and aligned in MUSCLE [55] with default settings. Phylogenetic inference was performed with BEAST version 10.5.0 using a 1+2 & 3 codon partition model with unique substitution rates and base frequencies for the partitions. Each partition was fitted to separate relaxed uncorrelated lognormal molecular clock models using random starting trees, four-category gamma-distributed HKY substitution models, and a birth-death process tree-shaped prior [58]. Four independent BEAST runs were performed, each with 100 million Markov chain Monte Carlo (MCMC) iterations sampled every 1,000 iterations. Runs were combined using LogCombiner with a burn-in of 20% and evaluated for convergence, sampling, and autocorrelation using Tracer version 1.7.2 [59]. A maximum clade credibility tree was inferred from the posterior sample and visualized with ggtree [60].

### Epidemiological model structure

An age-structured, stochastic SEIRS model was used to explore the population dynamics of co-circulating SARS-CoV-2 and SARS-CoV-X in the presence of vaccination (Supplementary Figure 1). Within this structure, susceptible (S) individuals may encounter infectious individuals with SARS-CoV-2 (I2) or SARS-CoV-X (IX) and transition to a corresponding exposed compartment (E2 or EX), with the likelihood of S-to-E transition controlled by the transmission probabilities of each virus. Exposed individuals then transition to an infectious state (I2 or IX), with the probability of E-to-I transition inferred from the incubation rate of each virus, and from infectious to recovered (R2 or RX) with a probability inferred from the recovery rates of each virus. Recovered individuals are considered immune to the virus they have recovered from but may transition back to a naïve state over time, with the R-to-S transition probability calculated from the waning rates of infection-acquired immunity.

To allow for re-infection of recovered individuals by the circulating SARS coronavirus, recovered individuals may also, upon encountering an infectious individual, transition to a separate exposed state (E2_RX_ or EX_R2_). These transitions occur with probabilities distinct from to those of S-to-E, allowing immunity from one virus to reduce susceptibility to infection with the other. Re-infected individuals then transition through separate infectious compartments (I2_RX_ or IX_R2_) – allowing re-infections to have distinct mortality and transition probabilities to infections of immunologically naïve individuals – before transitioning to the same recovered compartment (R_all_) where individuals are refractory to both viruses. Unique EIR compartments exist for each SARS-CoV-2 variant (Wuhan, Alpha, Delta, Omicron), allowing their infection phenotypes to vary, and additional E and I compartments exist for individuals recovered from a previous SARS-CoV-2 variant that have been re-infected with a subsequent variant. This allows recovered individuals who are immune to one variant (e.g., Wuhan) to be re-infected with a later variant, albeit with some cross-immunity.

Three distinct levels of this base structure were used to represent different vaccination statuses (unvaccinated, “U”; protected by 1 dose, “V1”; and protected by 2 doses, “V2”). Individuals in a lower level of vaccine protection may transition to their equivalent compartment in a higher level of vaccine protection with a probability based on the current population vaccination rate, and from a higher level to unvaccinated at a probability based on the rate of decay of SARS-CoV-2 vaccine-derived antibodies. By representing vaccination in this way, a distinction is made between the current protection phenotype of an individual, which increases with vaccination and decreases with waning immunity, and an individual’s vaccination history, where the number of doses may only increase with time. This allows an individual with a vaccination history of many doses to have an unprotected phenotype, provided enough time has passed from their last dose for the protection to have waned. Given the timing of vaccinations during and after the COVID-19 pandemic in the model population (Scotland), a negligible number of individuals achieved a protected by 3 doses (“V3”) phenotype throughout the model run, and so a fourth level representing these individuals was omitted for parsimony (Supplementary Figure 2).

### Host population structure

The population of Scotland was used as the basis for the host model population, for which there is detailed demographic and epidemiological information from before the COVID-19 pandemic to the present. Where information directly describing this population was unavailable, such as in the case of social contact data during and after the pandemic, information from the UK-wide population was used. Each compartment of the model was sub-divided into 16 age groups of 5 years, beginning 0-4 years old and ending 75+ years old, and the starting population sizes of each group were set to the projected population estimates from the National Records of Scotland for 2020. The birth rates of each age group were set as:

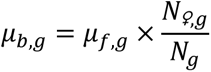

where *μ*_*b*,*g*_ is the birth rate, *μ*_*f*,*g*_ the fertility rate, *N_♀_*_,*g*_ the female population size, and *N*_*g*_ the total population size of age group *g*. All births were added to the 0-4 age group, and increases in age group occurred deterministically at a daily rate of 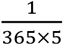. Age-stratified mortality and net migration rates were taken from publicly available data (see Supplementary Table 7 for all parameterisation sources). Migrants were assumed to have the same epidemiological properties as the Scotland population at the point of migration. For example, if 5% of the model population were in the I2 compartment, each migrant entering the model at that time point had a probability of 0.05 of being infectious for SARS-CoV-2. These parameters together created a host population that was largely stable over the 8-year duration of the model runs, with size and age progression similar to official population projections (Supplementary Figure 10).

To represent changes in social contacts over time, questionnaire data from multiple social mixing studies were used to calculate age-specific per-person per-day contact rates for different time periods. For the pre-lockdown period of 01/01/2020 (dd/mm/yyyy) to 23/03/2020, rates were set to those of the 2008 POLYMOD study of the UK population [61], with the study’s contact sub-categories (“home”, “work”, “school”, and “other”) summed to produce an overall daily rate of contact. For the period of 24/03/2020 to 02/03/2022, UK social contact information was collected continuously by the CoMix study [62]. In this study, the authors split their data into nine timeframes describing different lockdown and easing periods. To provide a finer temporal scale to the epidemiological model, this data was re-analysed to provide weekly estimates of contact rate using the same approach as the original study. Briefly, participant contacts were censored to a daily maximum of 50, and the mean number of contacts inferred from a negative binomial model of structure:

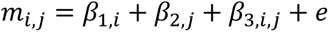

Here, *m*_*i*,*j*_ represents the mean number of contacts of participant age group *i* to contact age group *j*; *β*_1,*i*_ is the effect of participant group *i* on the mean number of contacts; *β*_2,*j*_ is the effect of contact group *j* on the mean number of contacts; *β*_3,*i*,*j*_ is the interaction effect of participant group *i* and contact group *j* on the mean number of contacts; and *e* is the model residuals. Symmetrical per-capita contact matrices were calculated from the output of this model using the following equation:

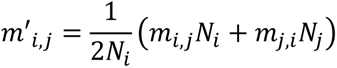

Here, *m*′_*i*,*j*_ is the adjusted mean number of contacts, *N*_*i*_ the total number of individuals in age group *i*, and *N*_*j*_ the total number of individuals in age group *j*. A follow-up to the CoMix study was conducted with data collected from 17/11/2022 to 07/12/2022, providing the most up-to-date estimates of UK contact rates available [63]. Data from this study was used to set the model contact rates from 17/11/2022 to the end of the model run (01/01/2028). Weekly contact rates in the inter-study period of 03/03/2022 to 16/11/2022 were inferred by linear interpolation from the final week of data of the original study to the first week of the follow-up study.

### Virus and immune phenotype parameterization

Parameter values for the infection-related mortality rates and incubation periods (E-to-I) of SARS-CoV and each SARS-CoV-2 variant were obtained from the literature (Supplementary Table 7). Literature estimates of the recovery rate (I-to-R) and *R*_0_for SARS-CoV-2 were highly variable and showed little consensus. Instead, these parameters were estimated by fitting the epidemiological model to prevalence data taken from the UK Coronavirus Infection Survey [64], using a grid search to minimize the sum of squared residuals. For SARS-CoV, literature estimates of *R*_0_were available, but no estimates were available for the recovery rate. Additionally, no information on the infection phenotypes of the prospective zoonotic SARS coronaviruses in humans exists. In the absence of this information, all missing phenotypes were assumed to equal those of the most closely related SARS coronavirus with a known phenotype. For example, the *R*_0_ of Rs4084 was set equal to SARS-CoV, while those of RaTG13 and GX/P1E were set equal to the SARS-CoV-2 Wuhan variant.

Transmission rates were calculated from the following *R*_0_ equation, derived from the SEIRS model structure:

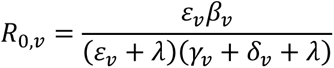

Here, *β*_*v*_ is the transmission rate; *R*_0,*v*_ the basic reproductive number; *ɛ*_*v*_ the incubation rate; *γ*_*v*_ the recovery rate; and *δ*_*v*_ the infection-related mortality rate of virus *v*, and *λ* is the population crude death rate. The transmission rates and infection-related mortality rates above describe the phenotypes of infection in naïve and unvaccinated individuals. Estimates of the proportion with which infection-related mortality is reduced in recovered and vaccinated individuals was taken from the literature. The proportion with which transmission rates are reduced in recovered and vaccinated individuals for SARS-CoV-X infections is assumed to correspond to the pseudotype neutralization data in Figure 1, and for SARS-CoV-2 re-infections from [53] (Supplementary Figure 9).

The waning rates of immunity in recovered individuals (R-to-S) and vaccinated individuals (V1-to-S and V2-to-S) were calculated separately from the half-lives of infection and vaccine derived antibodies against SARS-CoV-2 with the following exponential decay equation:

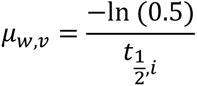

Here, *μ*_*w*,*v*_ represents the waning rate, and 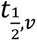 the antibody half-life of immunity of type *i* (natural or vaccine).

### Age-stratified phenotypes

There is strong evidence in the literature that immune protection and infection-related mortality for coronavirus infections vary across age groups. To represent this variation, non-linear least squares models, fitted using the nls function from the R core stats package [65], were used to infer how the relationship between phenotype and age changes with the population parameter value. For infection-related mortality, age-stratified data on SARS-CoV-2 and MERS-CoV were used to fit a four-parameter logistic function (Supplementary Figure 11) as:

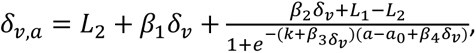

Here, *δ*_*v*,*a*_ is the infection-related mortality rate of virus *v* for hosts of age *a*; *k* is the slope, *L*_1_ the upper plateau, and *L*_2_ the lower plateau of the logistic function; and *a*_0_ is the inflection point. *β*_1_*δ*_*v*_, *β*_2_*δ*_*v*_, *β*_3_*δ*_*v*_, and *β*_4_*δ*_*v*_ describe how the population infection-related mortality rate affects the lower plateau, upper plateau, slope, and midpoint of the logistic function respectively.

The effectiveness of the second dose of the COVID-19 vaccine, administered at different intervals from the first dose, was used to provide a dataset where the population-level estimate of immune protection varied, and the age-stratified estimates of immune protection were known (Supplementary Figure 12). This data was used to fit a model with a formula composed of two three-parameter logistic functions (one describing the increase in immune effectiveness from birth to middle age, the other describing the decline in effectiveness from middle age to old age), and an intercept influenced by the population-level protection:

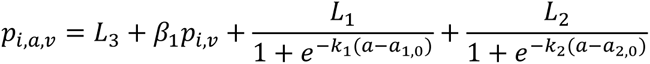

Here, *p*_*i*,*a*_ is the level of protection conferred by immunity source *i* against virus *v* on age group *a*. *L*_1_, *k*_1_, and *a*_1,*o*_ are the upper plateau, slope, and infection point of the increase in immune effectiveness with age, while *L*_2_, *k*_2_, and *a*_2,*o*_ are the equivalents for the decrease in immune effectiveness with age. *β*_1_*p*_*i*,*v*_ describes the effect of population-level immune protection on the function’s intercept (*L*_3_). Little conclusive evidence exists for an effect of host age on the incubation or recovery rates of SARS coronavirus infections [66–70], and so these parameters are assumed to be equal across age groups.

### Vaccination rates

For the initial COVID-19 vaccination program (08/12/2020 to 11/09/2022), daily vaccination rates for each age group receiving the first (*u*_*D*1_), second (*u*_*D*2_), and third (*u*_*D*3_) vaccine doses were calculated from publicly available data [71]. As vaccination exists in the epidemiological model as a transient phenotype with three levels (U, V1, V2), two distinct rates exist: the transition rate from naïve to protected with 1 dose (*u*_*U*→*V*1_) and the transition rate from protected with 1 dose to protected with 2 doses (*u*_*V*1→*V*2_). As entry into the V2 level is impossible while only 1 dose of the vaccine has been administered, this rate was set as *u*_*V*1→*V*2_ = *u*_*D*2_ + *u*_*D*3_ . Transitioning from a naïve to vaccinated phenotype can be produced at any point in the vaccination program and by any dose in a patient’s vaccination history, and so this rate was set as *u*_*U*→*V*1_ = *u*_*D*1_ + *u*_*D*2_ + *u*_*D*3_ . Vaccination rates by age group during the booster programs in Winter 2022, Spring 2023, Winter 2023, and Spring 2024 were used to extend *u*_*D*3_ to 14/07/2024 [72]. Spring and Winter booster programs were then assumed to recur annually at rates identical to Winter 2023 and Spring 2024 for the remainder of the model.

The rate of administration of the first COVID-19 vaccine dose over time in Scotland is well approximated by a non-normalised Gaussian curve (Supplementary Figure 13), and so a similar curve with a mean (µ) of 30 days and standard deviation (σ) of 10 days was used to define a theoretical preventative campaign against SARS-CoV-X emergence lasting 60 days:

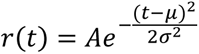

By adjusting the scaling factor (A), the vaccination rates described by this curve result in different cumulative vaccine uptake levels over the course of the campaign. Solutions for the value of A that produces specific values of vaccine uptake were calculated using root-finding methods with the uniroot function in R [65].

### Stochasticity

Mean rates for all model parameters (excluding the aging rate) were converted to probabilities using a Euler scheme with binomial increments and exponential rates [73] such that the number of individuals *N* transitioning from compartment *A* to *B* at time *t* is given as:

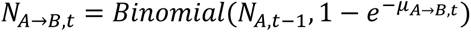

Here, *Binomial*(*n*, *p*) represents the binomial distribution, *n* is the number of individuals available to a given transition, and *p* is the probability of transition calculated from the mean rate *u*.

### Model run conditions

The model was implemented and run using the Odin [74] and Odin Dust [75] R packages, with random starting seeds, a 1-day time step, and a duration of 8 years spanning 1^st^ January 2020 to 1^st^ January 2028. To introduce the SARS-CoV-2 Wuhan variant, five exposed individuals (E2_Wuhan_) were added to the model on 23^rd^ February 2020. This date was inferred from the description of the first confirmed positive case of SARS-CoV-2 infection in Scotland [76], which was reported on 2^nd^ March 2020 in an individual presenting 3 days after symptom onset (-3 days) and after a ∼5-day incubation period (-5 days). Five individuals were chosen as this ensured SARS-CoV-2 reached endemicity in all model runs conducted in this study. Introduction of new SARS-CoV-2 variants to the model occurred deterministically by taking the total number of SARS-CoV-2 infectious individuals, regardless of variant, and multiplying by the prevalence of each variant as reported for the Scotland population over time. These values were then used in conjunction with contact probabilities to calculate the number of individuals exposed to each variant as the model progressed. A single SARS-CoV-X individual (E_X_) was introduced to the model on 23^rd^ February 2024, four years after the initial SARS-CoV-2 exposure.

## Data Availability

All data produced are available online from GitHub (https://github.com/ryanmimrie/Publications_2025_SARS-CoV-X-Emergence) and FigShare (https://doi.org/10.6084/m9.figshare.28566677.v1).

https://doi.org/10.6084/m9.figshare.28566677.v1

## Author contributions

Conceptualization: MV, BJW, PRM

Methodology: RI, MV, BJW, LM, MB, PRM

Investigation: RI, LAB, SR, MM, JARA, LM, NL

Visualization: RI, LAB, MM, JARA

Funding acquisition: PRM, BJW, MV

Project administration: PRM, BJW

Supervision: MV, BJW, PRM

Writing – original draft: RI, LAB, MV, BJW, PRM

Writing – review & editing: RI, LAB, SR, MM, JARA, LM, NL, AP, MB, MV, BJW, PRM

## Data and materials availability

All data and example modelling scripts used in this study can be found on GitHub (https://github.com/ryanmimrie/Publications_2025_SARS-CoV-X-Emergence) and FigShare (https://doi.org/10.6084/m9.figshare.28566677.v1).

