## Supplementary Text for "Post-pandemic changes in population immunity have reduced the likelihood of emergence of zoonotic coronaviruses"

**This file includes:**

- Supplementary Figures
- Supplementary Tables

**Supplementary Figures:**

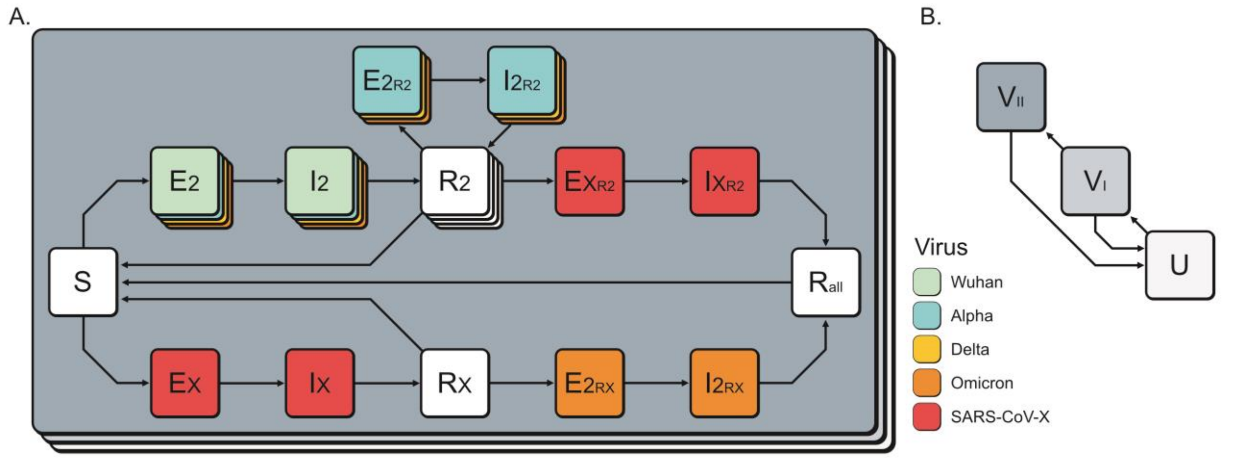


**Supplementary Figure 1: Epidemiological model schematic representing the transmission dynamics of co-circulating SARS-CoV-2 and SARS-CoV-X in the presence of vaccination.** The core structure of the model, represented in (A.), contains five unique EI compartment groups describing i) infection of naïve (S) individuals with SARS-CoV-2 (E2, I2); ii) infection of naïve individuals with SARS-CoV-X (EX, IX); iii) re-infection of individuals recovered from an earlier variant of SARS-CoV-2 (R2) with a later variant ($E2_{R2}$, $I2_{R2}$); iv) re-infection of individuals recovered from SARS-CoV-2 with SARS-CoV-X ($EX_{R2}$ and $IX_{R2}$); and v) re-infection of individuals recovered from SARS-CoV-X (RX) with SARS-CoV-2 ($E2_{RX}$ and $I2_{RX}$). This structure is repeated across three layers representing different levels of vaccine protection against infection (B.), where individuals may be unvaccinated (U), protected by a single dose ($V_I$), or protected by two doses ($V_{II}$).

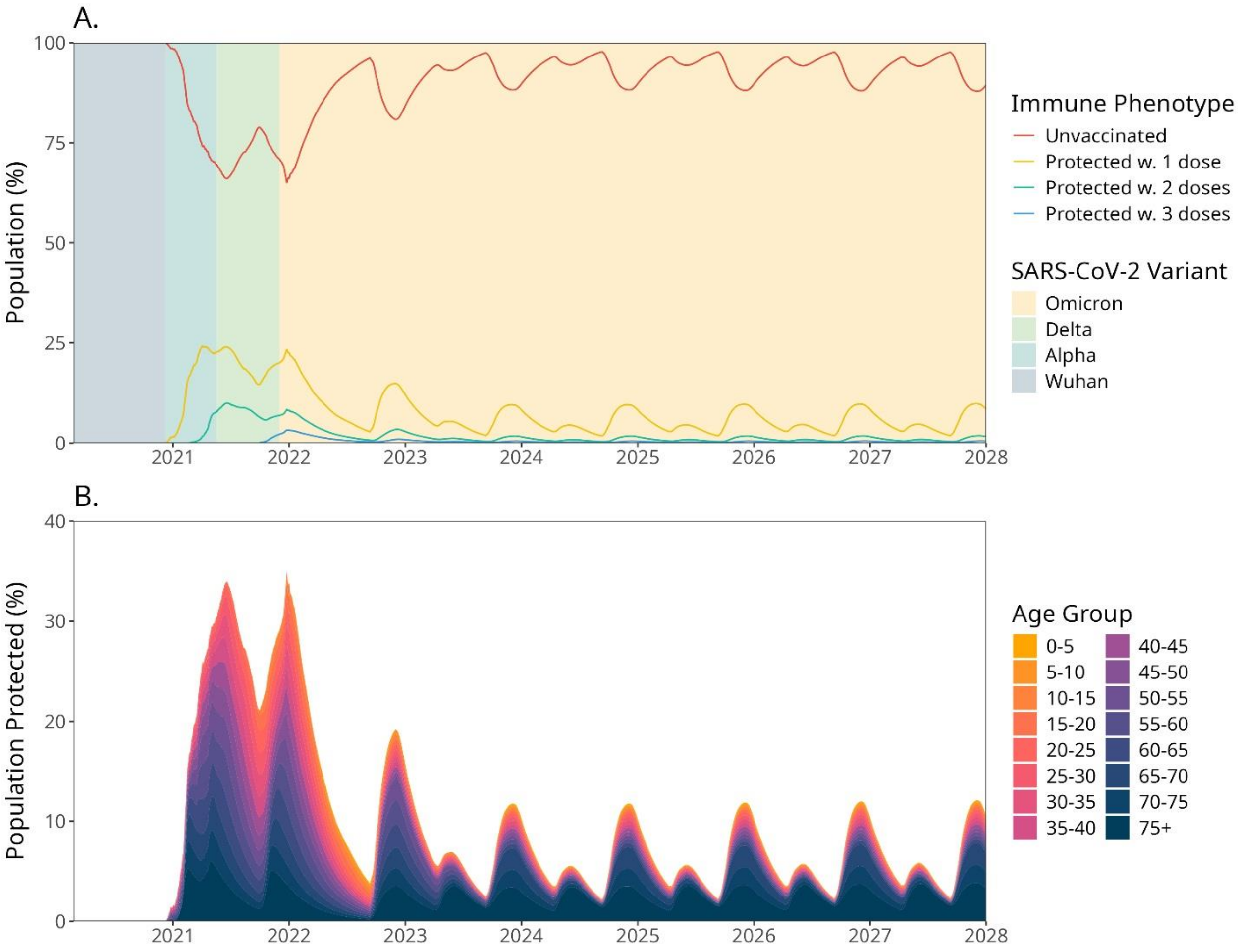


**Supplementary Figure 2: Population coverage of vaccination protection over time.** A) Lines show the number of individuals with an unvaccinated (red), protected with one dose (yellow), two doses (green), or three doses (blue) immune phenotype as a percentage of the total population over time, in an example model run under similar conditions to Figure 2A, but with an additional vaccination level. The predominant SARS-CoV-2 variant is indicated by the shaded background area for reference. B) The percentage of individuals in the population protected by vaccination over time in the three-level (unvaccinated, 1 dose, 2 dose) model used throughout this study, separated by age group.

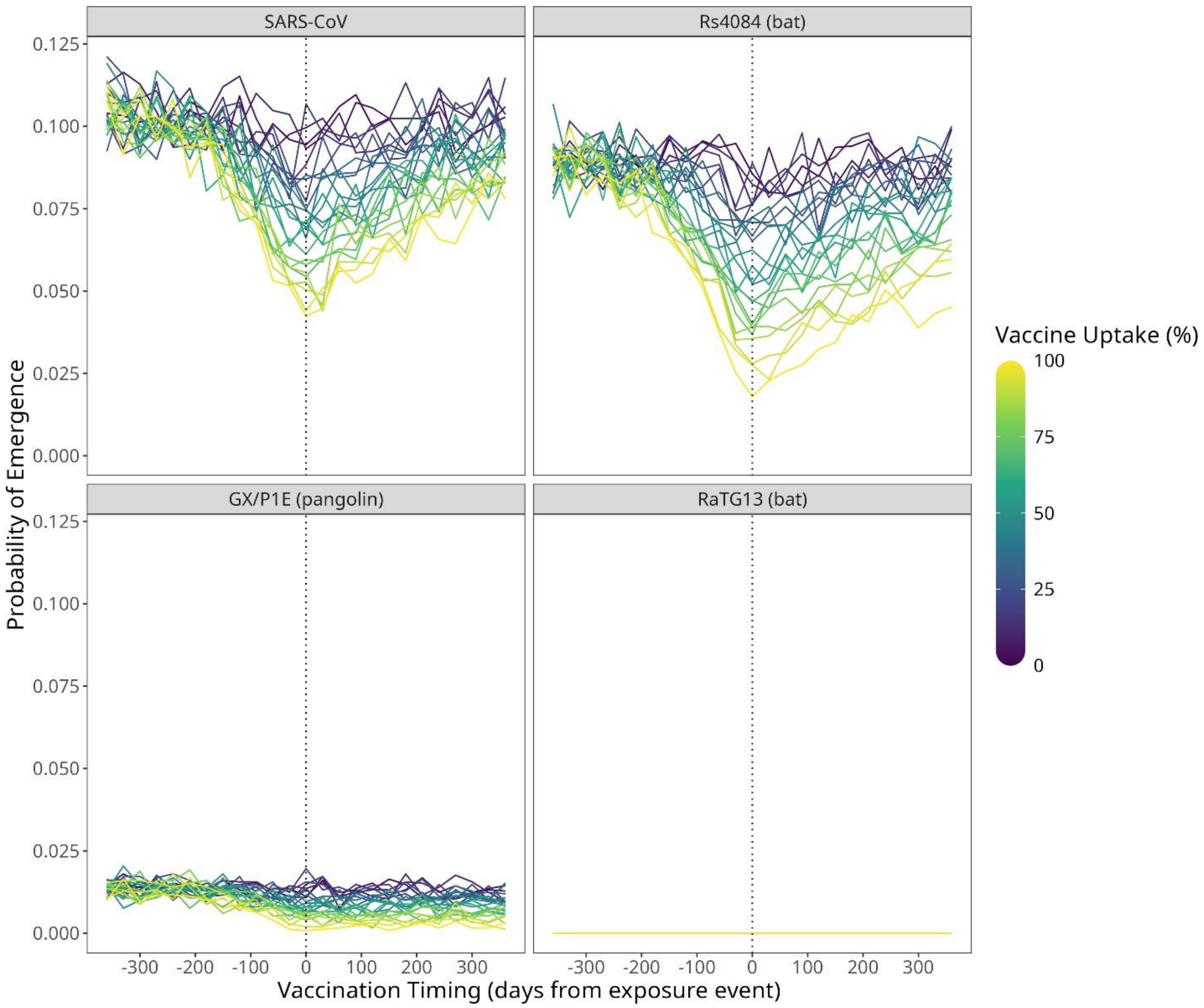


**Supplementary Figure 3: Probability of emergence of different SARS coronaviruses in the presence of vaccination and co-circulating SARS-CoV-2.** Lines represent the point estimates of the probability of emergence (y-axis) for four SARS coronaviruses in a population with co-circulating SARS-CoV-2 under preventative vaccination programs at different times relative to the SARS-CoV-X exposure event (x). The colour of each line indicates the (%) uptake of the vaccination program.

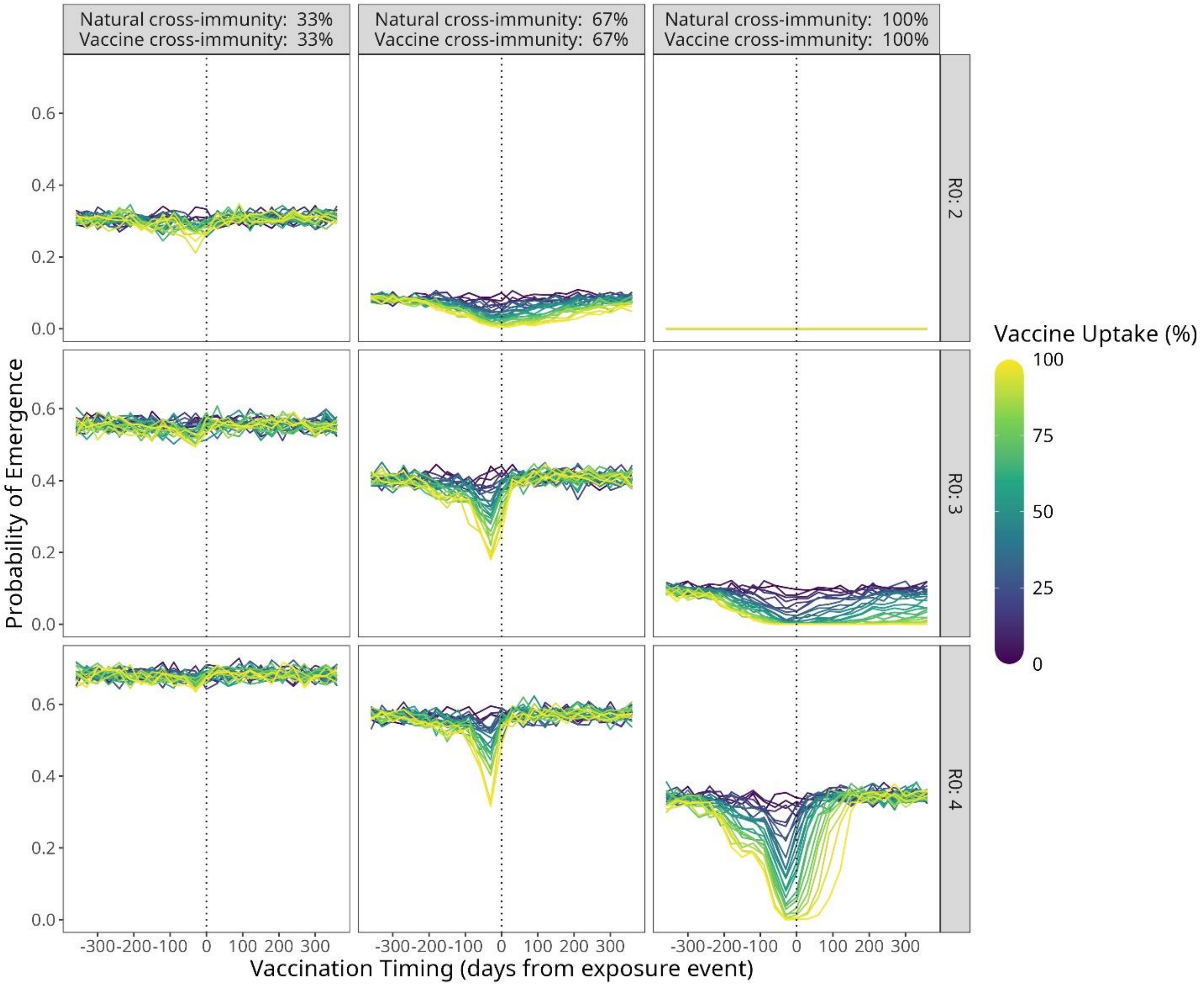


**Supplementary Figure 4: Probability of emergence of theoretical SARS coronaviruses under conditions of equal cross-immunity and vaccine effectiveness.** Lines show point estimates of the probability of emergence for nine theoretical SARS coronaviruses with different R0 values (facet rows) and conditions of cross-immunity and vaccine effectiveness (facet columns) in a population with co-circulating SARS-CoV-2. In these scenarios, protection against SARS-CoV-X infection conferred from recovering from natural infection with SARS-CoV-2 (“natural cross-immunity”) and vaccination (“vaccine cross-immunity”) are identical.

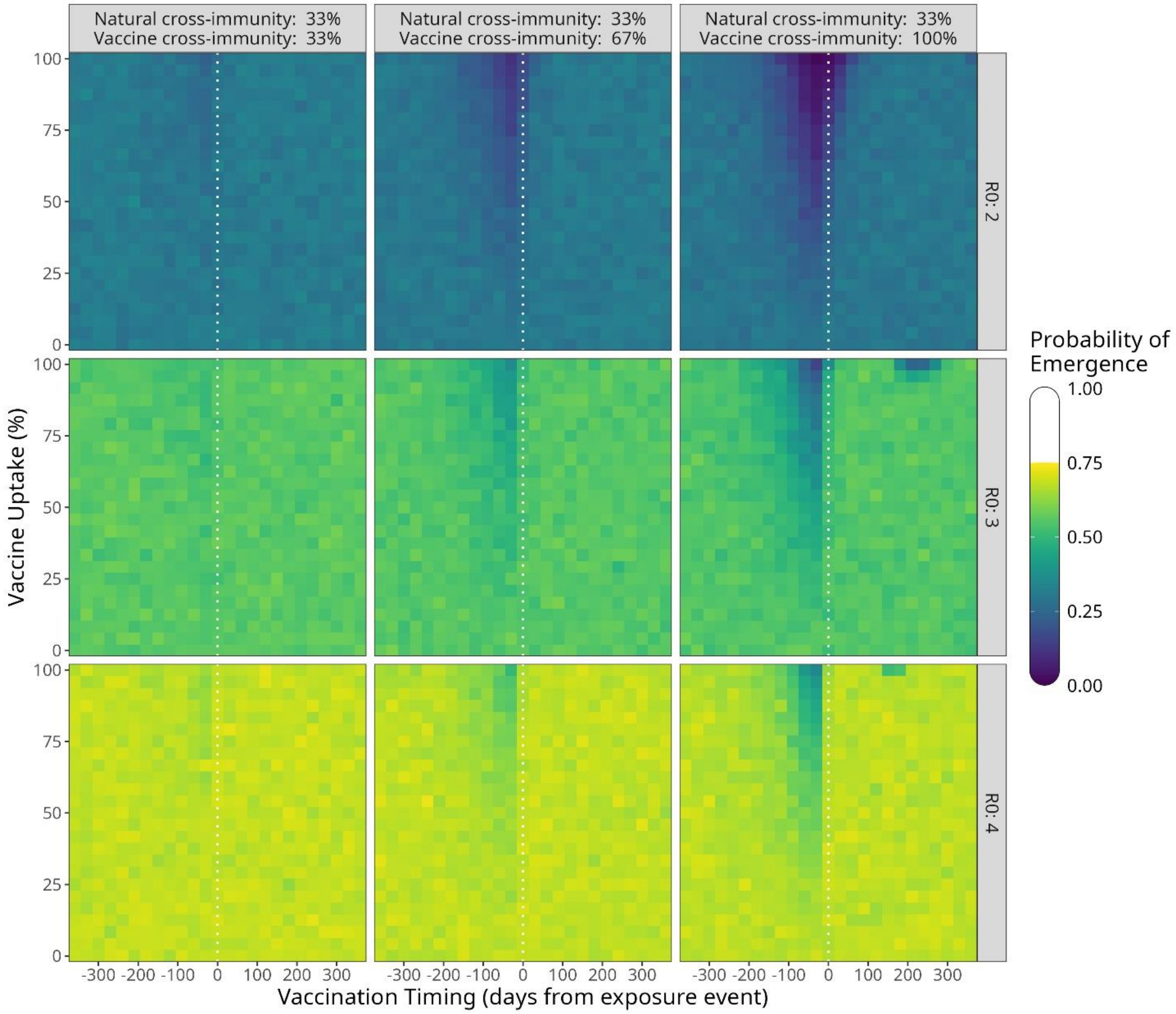


**Supplementary Figure 5: Probability of emergence of theoretical SARS coronaviruses under conditions of varying vaccine cross-immunity.** Heatmaps show point estimates of the probability of emergence for nine theoretical SARS coronaviruses with different R0 values (facet rows) and varying conditions of vaccine cross-immunity (facet columns) in a population with co-circulating SARS-CoV-2.

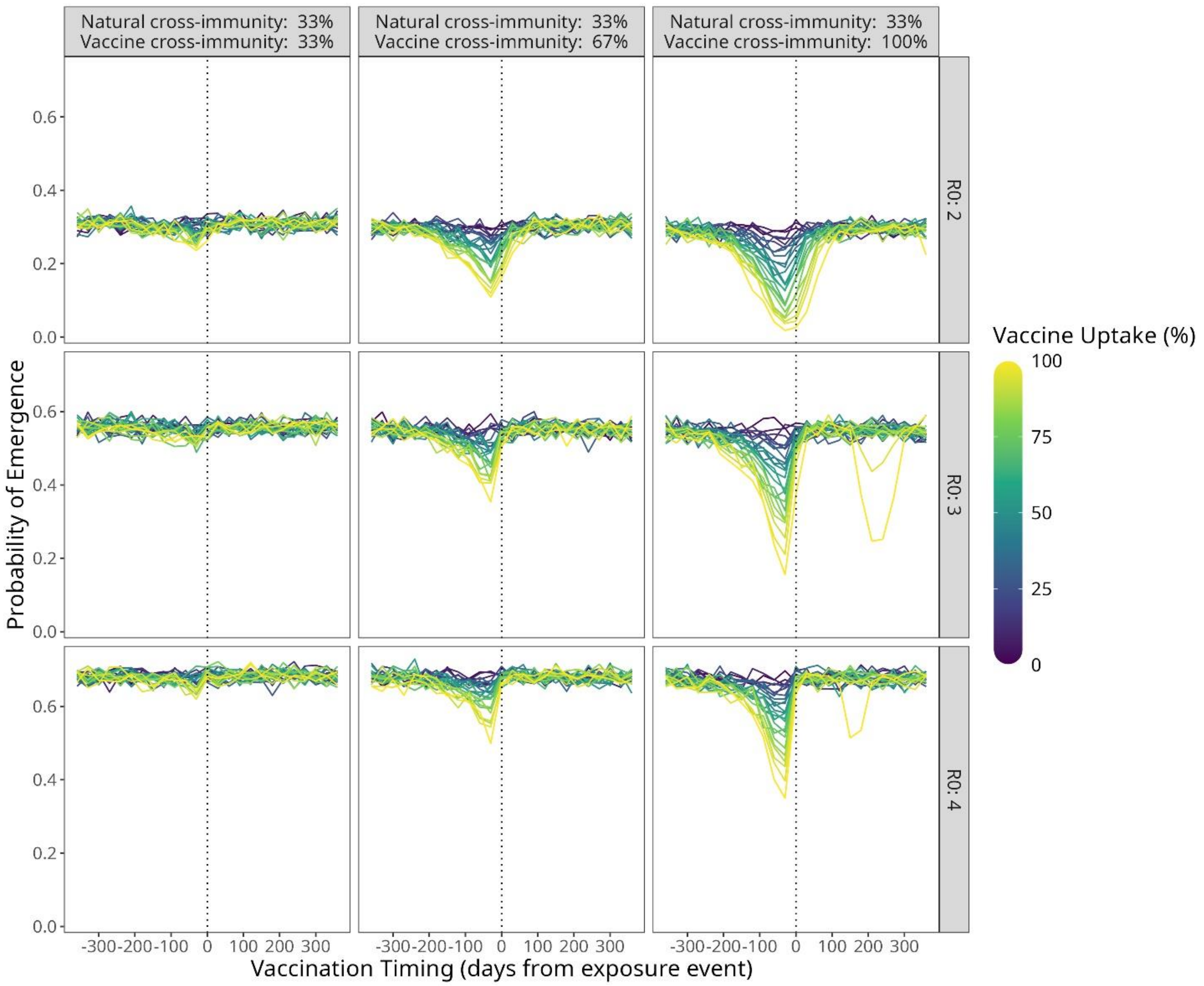


**Supplementary Figure 6: Probability of emergence of theoretical SARS coronaviruses under conditions of varying vaccine effectiveness.** Lines show point estimates of the probability of emergence for nine theoretical SARS coronaviruses with different R0 values (facet rows) under varying conditions of vaccine effectiveness (facet columns), in a population with co-circulating SARS-CoV-2.

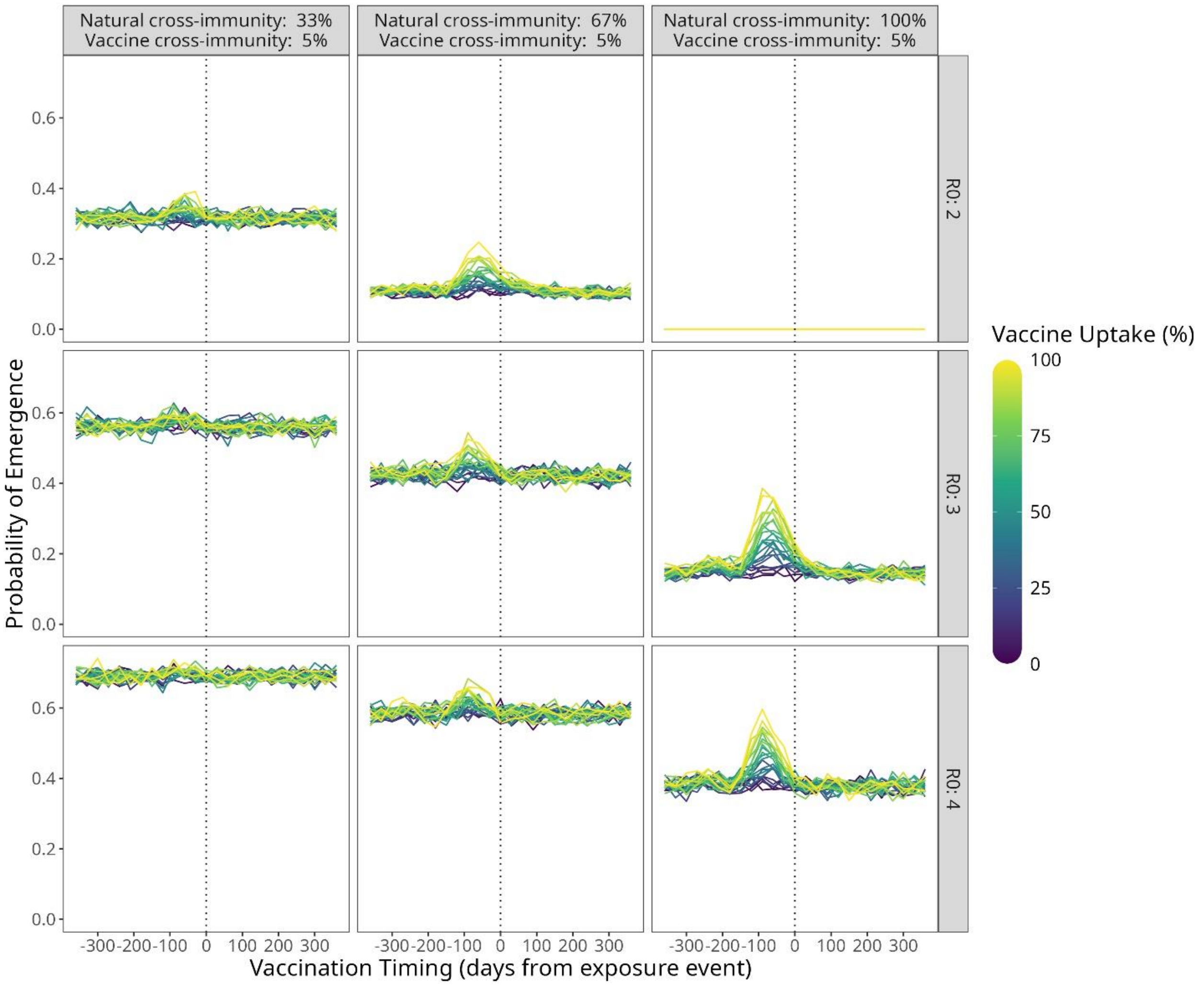


**Supplementary Figure 7: Probability of emergence of theoretical SARS coronaviruses under conditions of low vaccine cross-immunity and high natural cross-immunity.** Lines show point estimates of the probability of emergence for nine theoretical SARS coronaviruses with different R0 values (facet rows) and varying conditions of natural cross-immunity (facet columns) in a population with co-circulating SARS-CoV-2 and low (5%) vaccine cross-immunity.

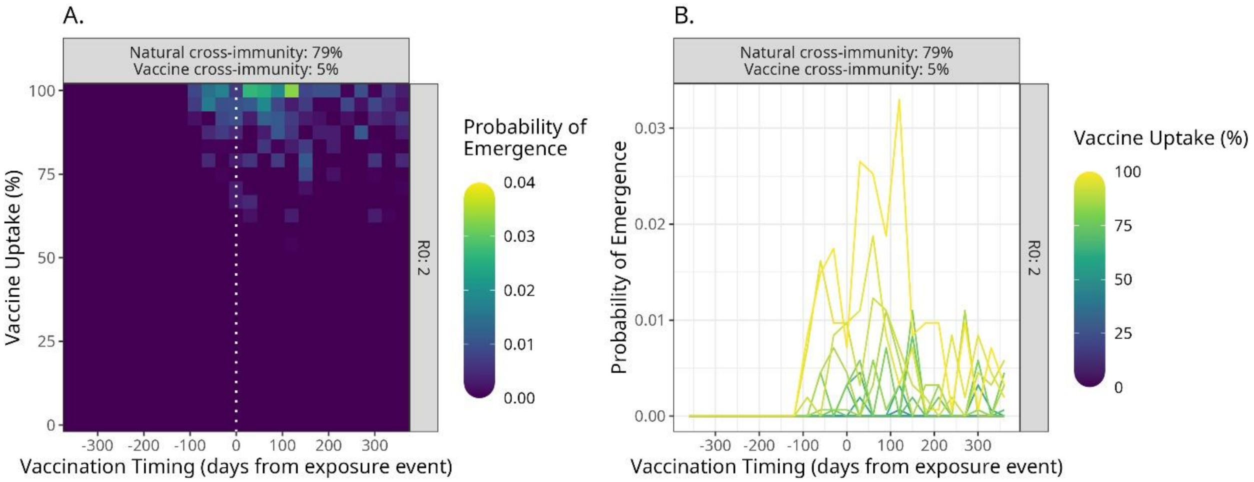


**Supplementary Figure 8: Probability of emergence of a theoretical SARS coronavirus under conditions of low vaccine cross-immunity and natural cross-immunity of 79%.** The heatmap shows point estimates of the probability of emergence for a theoretical SARS coronavirus with R0 = 2 in a population with co-circulating SARS-CoV-2, under conditions of 79% natural cross-immunity and 5% vaccine cross-immunity.

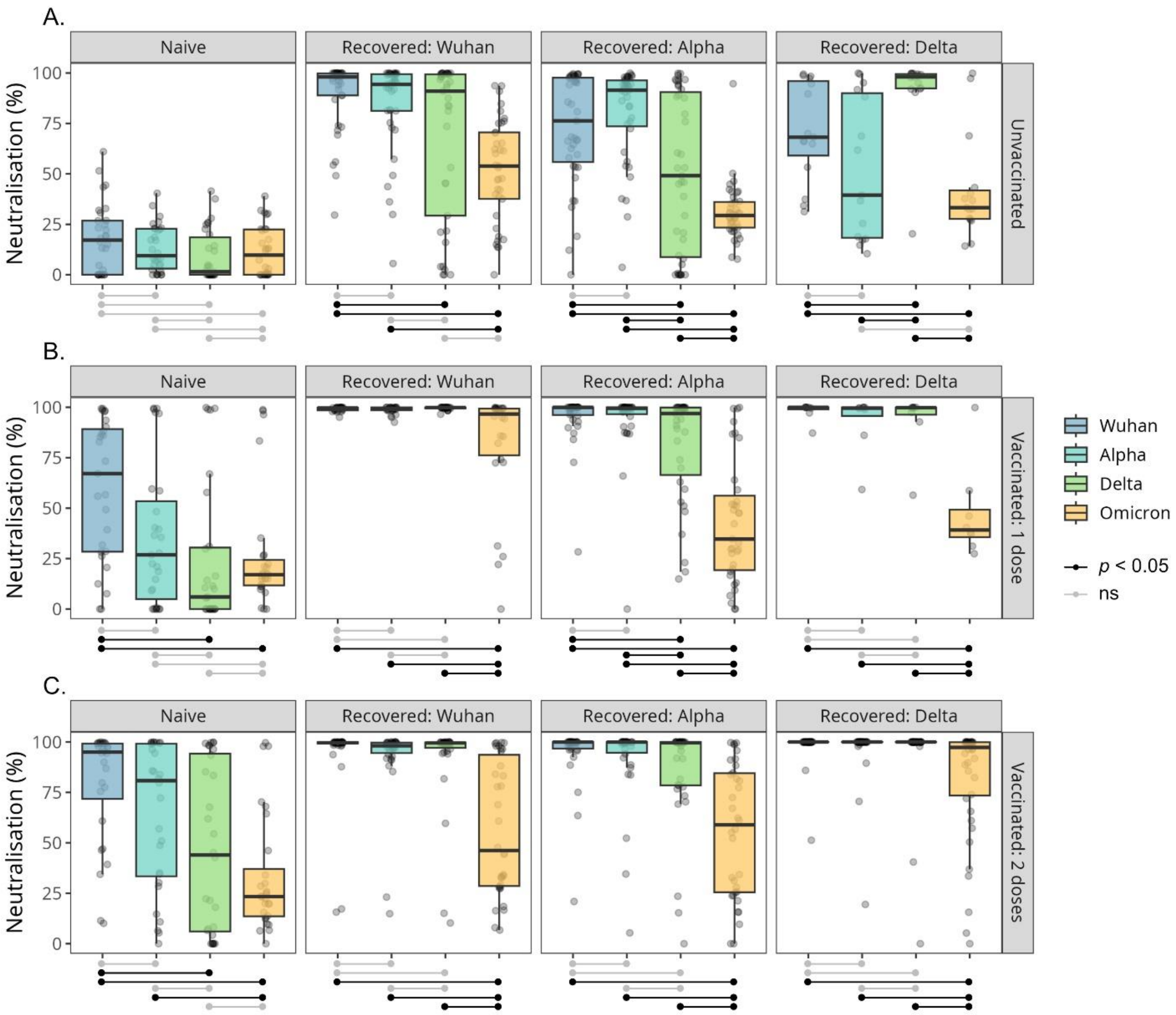


**Supplementary Figure 9: Neutralisation of viral pseudotypes carrying the spike proteins of different SARS-CoV-2 variants by sera from individuals of different infection and vaccination histories.** Boxplots show the percentage neutralisation of pseudotype viruses by sera from individuals who were unvaccinated (A), vaccinated once (B), or vaccinated twice (C) against SARS-CoV-2. Results are separated into subplots (columns) based on an individual's history of natural infection, and separate boxplots are shown for neutralisation of pseudoviruses with Wuhan (blue), Alpha (cyan), Delta (green) and Omicron (yellow) spike proteins. Significant differences in the strength of neutralisation are shown with black horizontal lines below each subplot, assessed using Welch's t-tests with Holm correction for multiple testing.

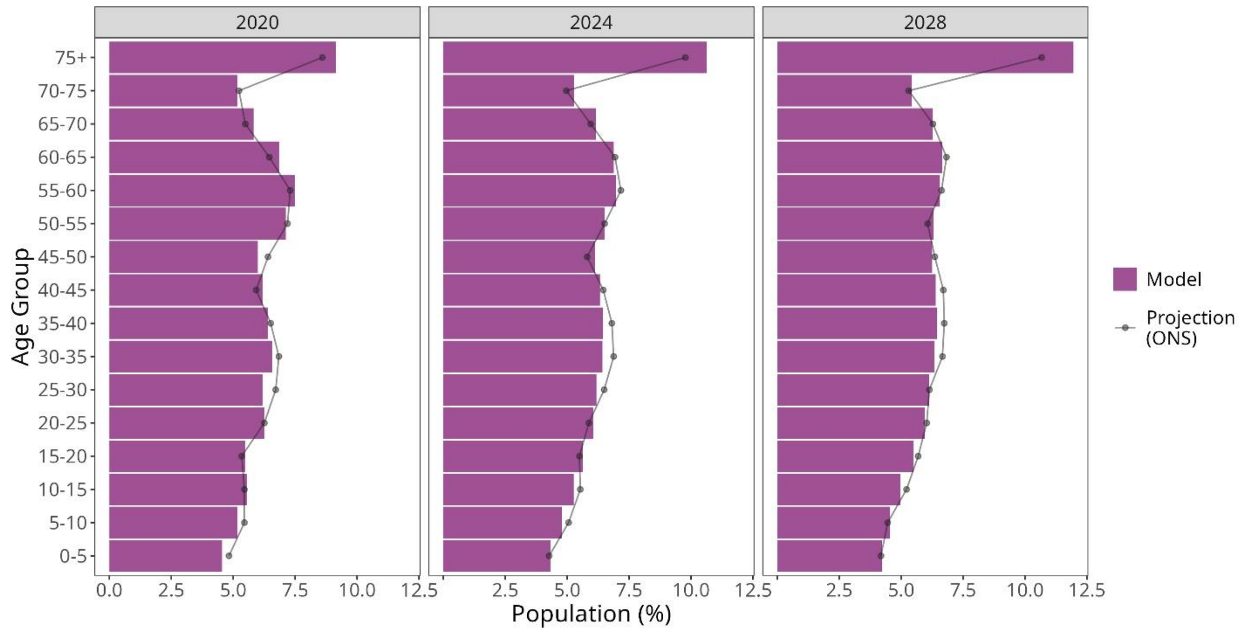


**Supplementary Figure 10: Population percentages by age-group over time.** Orange bars show the number of individuals in each age group as a percentage of the total population on January 1st 2020, 2024, and 2028. Grey lines indicate the population projections for Scotland estimated by the Office for National Statistics (ONS), available from: https://www.nrscotland.gov.uk/statistics-and-data/population-migration-and-households/

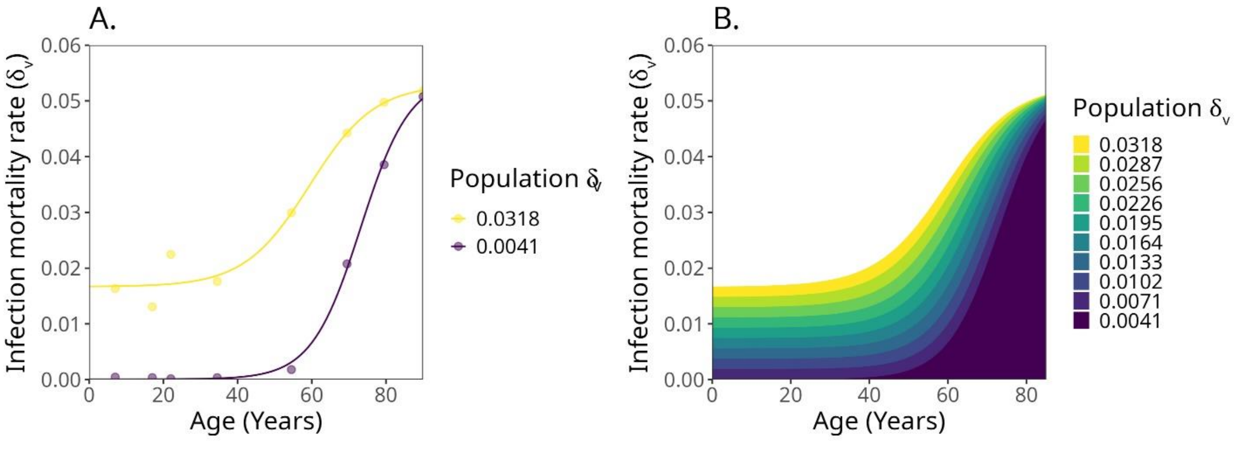


**Supplementary Figure 11: Inferring infection mortality rate across age-groups for theoretical SARS coronaviruses of different population infection mortality rates.** A) Infection mortality rates of different ages for SARS-CoV-2 (purple) and MERS-CoV (yellow) with a four-parameter logistic non-linear least squares model fitted to each group. B) Using the model fit from (A), infection mortality rates of different ages are inferred for viruses with different population infection mortality rates.

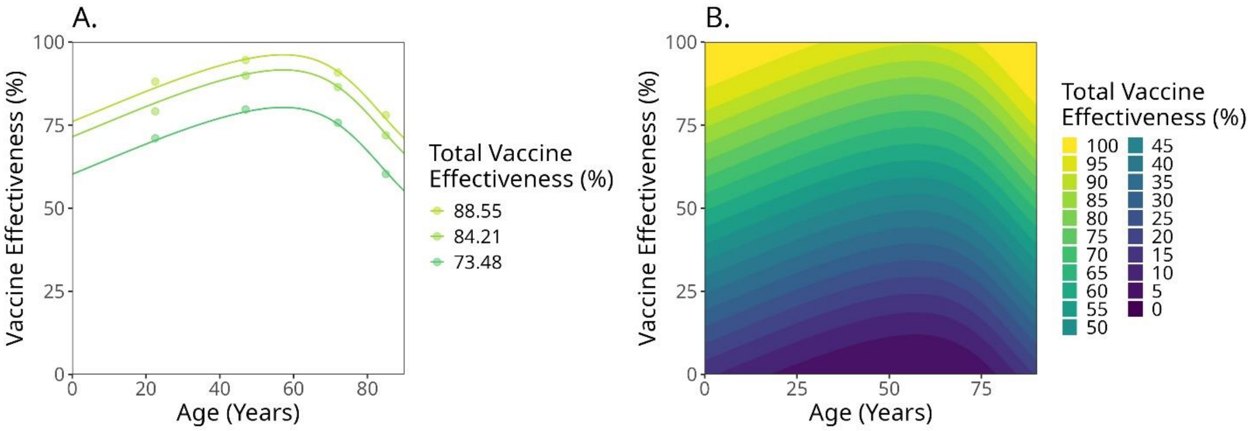


**Supplementary Figure 12: Inferring the levels of immune protection across age groups for different levels of population protection.** A) Estimates of COVID-19 vaccine effectiveness against re-infection for different ages for those who received a 2nd dose within 3 months (light green), 3-6 months (mid green), or later than 6 months (dark green) from their 1st dose. A double-logistic non-linear least squares model is fitted to each group. B) Using the model fit from (A), the levels of immune protection across different ages are inferred for sources of immunity with different levels of population protection.

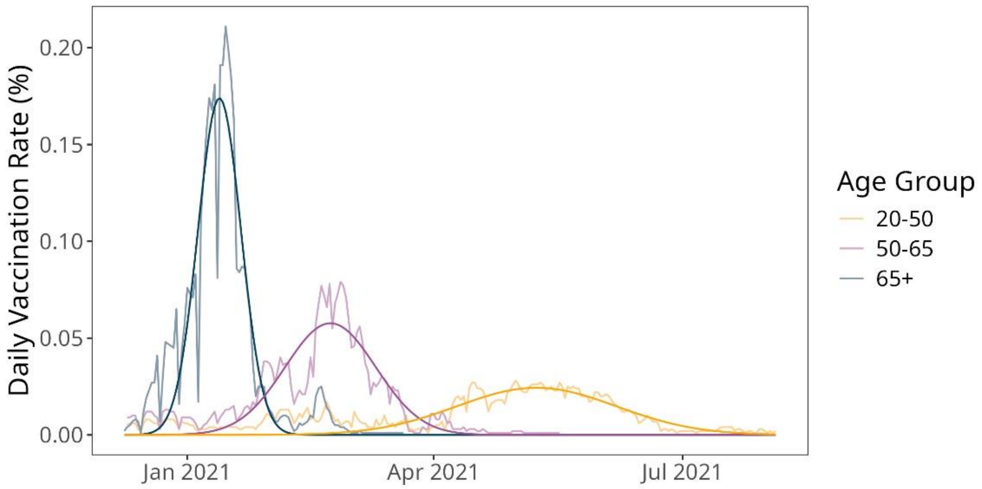


**Supplementary Figure 13: The rate of administration of the first COVID-19 vaccine dose in Scotland**. Data on the daily rate of vaccination for the first COVID-19 dose (faded lines) is shown for age groups 65+ (dark blue), 50-65 (purple), and 20-50 (yellow). Non-normalised Gaussian curves, fitted using a non-linear least squares approach, are overlaid in darker lines.

### Supplementary Tables:

**Supplementary Table 1: Across-viruses mixed effect model random effects.**

| Group | Variance | Std. Deviation |
|---|---|---|
| Serum.ID | 296.3 | 17.21 |
| Virus | 451.2 | 21.24 |
| Residual | 358.7 | 18.94 |

**Supplementary Table 2: Across-viruses mixed effect model fixed effects.** Intercept = Naïve & Unvaccinated

| Effect | Estimate (µ) | Std. Error | df | t-value | p-value |
|---|---|---|---|---|---|
| (Intercept) | 14.820 | 11.211 | 3.706 | 1.322 | 0.2619 |
| Recovered | 26.877 | 4.148 | 344 | 6.480 | <0.001 |
| Vaccinated 1 dose | 30.347 | 5.212 | 344 | 5.823 | <0.001 |
| Vaccinated 2 doses | 39.677 | 5.162 | 344 | 7.686 | <0.001 |
| Recovered : 1 dose | -7.918 | 6.090 | 344 | -1.300 | 0.1944 |
| Recovered : 2 doses | -14.348 | 5.882 | 344 | -2.439 | 0.0152 |

**Supplementary Table 3: Across-immune group mixed effect model random effects.**

| Group | Variance | Std. Deviation |
|---|---|---|
| Serum.ID | 296.45 | 17.218 |
| Recovered | 181.55 | 13.474 |
| Vaccinated | 265.05 | 16.280 |
| Recovered : Vaccinated | 15.72 | 3.965 |
| Residual | 358.65 | 18.938 |

**Supplementary Table 4: Across-immune group mixed effect model fixed effects.** Intercept = SARS-CoV-1

| Effect | Estimate (µ) | Std. Error | df | t-value | p-value |
|---|---|---|---|---|---|
| (Intercept) | 29.966 | 13.565 | 2.553 | 2.209 | 0.13 |
| Rs4084 | 9.761 | 1.432 | 1047 | 6.818 | <0.001 |
| GX/P1E | 13.349 | 1.432 | 1047 | 9.325 | <0.001 |
| RaTG13 | 48.713 | 1.432 | 1047 | 34.028 | <0.001 |

**Supplementary Table 5: Across-immune group mixed effect model random effects.**

| Group | Variance | Std. Deviation |
|---|---|---|
| Serum.ID | 267.39 | 16.352 |
| Recovered | 52.44 | 5.914 |
| Vaccinated | 163.26 | 7.241 |
| Recovered : Vaccinated | 34.98 | 12.777 |
| Residual | 433.28 | 20.815 |

**Supplementary Table 6: Across-immune group mixed effect model fixed effects.**

| Effect | Estimate (μ) | Std. Error | df | t-value | p-value |
|---|---|---|---|---|---|
| (Intercept) | 37.49292 | 8.52023 | 2.58108 | 4.40 | 0.0295 |
| Similarity^2 | 0.09249 | 0.00299 | 958.99953 | 30.93 | <0.001 |

**Supplementary Table 7: Parameterization Values and References.** Values are given as quantities (population sizes), daily rates (migration) or daily per-capita rates (all other values).

| Parameter | Population/Virus | Value(s) | | Reference(s) |
|---|---|---|---|---|
| *Population Size* | Scotland (by age) | 0-4: | 247737 | [1] |
| | | 5-9: | 281912 | |
| | | 10-14: | 303266 | |
| | | 15-19: | 298467 | |
| | | 20-24: | 341371 | |
| | | 25-29: | 337679 | |
| | | 30-34: | 357840 | |
| | | 35-39: | 348532 | |
| | | 40-44: | 336578 | |
| | | 45-49: | 326504 | |
| | | 50-54: | 388254 | |
| | | 55-59: | 408491 | |
| | | 60-64: | 373674 | |
| | | 65-69: | 317469 | |
| | | 70-74: | 282016 | |
| | | 75+: | 497910 | |
| *Birth Rate* | Scotland (by age) | 0-4: | 0 | [2] |
| | | 5-9: | 0 | |
| | | 10-14: | 0.0000002444 | |
| | | 15-19: | 0.0000166204 | |
| | | 20-24: | 0.0000478186 | |
| | | 25-29: | 0.0000952367 | |
| | | 30-34: | 0.0001204575 | |
| | | 35-39: | 0.0000601418 | |
| | | 40-44: | 0.0000110273 | |
| | | 45-49: | 0.0000005861 | |
| | | 50-54: | 0.0000000335 | |
| | | 55-59: | 0 | |
| | | 60-64: | 0 | |
| | | 65-69: | 0 | |
| | | 70-74: | 0 | |
| | | 75+: | 0 | |
| *Crude Death Rate* | Scotland (by age) | 0-4: | 0.00000187412 | [2] |
| | | 5-9: | 0.00000023801 | |
| | | 10-14: | 0.00000047469 | |
| | | 15-19: | 0.00000111622 | |
| | | 20-24: | 0.00000154121 | |
| | | 25-29: | 0.00000211211 | |
| | | 30-34: | 0.00000305548 | |
| | | 35-39: | 0.00000481195 | |
| | | 40-44: | 0.00000682835 | |
| | | 45-49: | 0.00000915344 | |
| | | 50-54: | 0.00001264592 | |
| | | 55-59: | 0.00001855913 | |
| | | 60-64: | 0.00002881181 | |
| | | 65-69: | 0.00004558282 | |
| | | 70-74: | 0.00007269524 | |
| | | 75+: | 0.00018167223 | |
| *Net Migration Rate* | Scotland (by age) | 0-4: | 3 | [1] |
| | | 5-9: | 3 | |
| | | 10-14: | 2 | |
| | | 15-19: | 14 | |

| | | | |
|---|---|---|---|
| | | 20-24: 13<br>25-29: 2<br>30-34: 5<br>35-39: 3<br>40-44: 2<br>45-49: 2<br>50-54: 2<br>55-59: 2<br>60-64: 2<br>65-69: 1<br>70-74: 0<br>75+: 0 | |
| *Contact Rates* | Scotland (by age) | Multi-dimensional. Available at: https://github.com/ryanmimrie/Publications_2025_SARS-CoV-X-Emergence | [3–5] |
| *Vaccination Rates* | Scotland (by age) | Multi-dimensional. Available at: https://github.com/ryanmimrie/Publications_2025_SARS-CoV-X-Emergence | [6,7] |
| *Vaccination Waning Rate* | Scotland | 0.01131 | Inferred through exponential decay model fitted to data in [8] |
| *SARS-CoV-2 Variant Prevalences* | Scotland | Multi-dimensional. Available at: https://github.com/ryanmimrie/Publications_2025_SARS-CoV-X-Emergence | [9] |
| *$R_0$* | SARS-CoV-2 (Wuhan) | 1.575 | Inferred through epidemiological model fitted to data in |
| | SARS-CoV-2 (Alpha) | 1.098 | Inferred through epidemiological model fitted to data in |
| | SARS-CoV-2 (Delta) | 1.697 | Inferred through epidemiological model fitted to data in |
| | SARS-CoV-2 (Omicron) | 2.511 | Inferred through epidemiological model fitted to data in |
| | SARS-CoV-1 | 1.575 | Assumed equal to SARS-CoV-2 (Wuhan) |
| | Rs4084 | 1.575 | Assumed equal to SARS-CoV-2 (Wuhan) |
| | GX/P1E | 1.575 | Assumed equal to SARS-CoV-2 (Wuhan) |
| | RaTG13 | 1.575 | Assumed equal to SARS-CoV-2 (Wuhan) |
| *Incubation Rate* | SARS-CoV-2 (Wuhan) | 0.217 | [10] |
| | SARS-CoV-2 (Alpha) | 0.202 | [10] |
| | SARS-CoV-2 (Delta) | 0.226 | [10] |
| | SARS-CoV-2 (Omicron) | 0.277 | [10] |
| | SARS-CoV-1 | 0.217 | Assumed equal to SARS-CoV-2 (Wuhan) |
| | Rs4084 | 0.217 | Assumed equal to SARS-CoV-2 (Wuhan) |
| | GX/P1E | 0.217 | Assumed equal to SARS-CoV-2 (Wuhan) |
| | RaTG13 | 0.217 | Assumed equal to SARS-CoV-2 (Wuhan) |
| *Recovery Rate* | SARS-CoV-2 (Wuhan) | 0.048 | Inferred through epidemiological model fitted to data in [11] |
| | SARS-CoV-2 (Alpha) | 0.119 | Inferred through epidemiological model fitted to data in [11] |
| | SARS-CoV-2 (Delta) | 0.153 | Inferred through epidemiological model fitted to data in [11] |
| | SARS-CoV-2 (Omicron) | 0.275 | Inferred through epidemiological model fitted to data in [11] |
| | SARS-CoV-1 | 0.048 | Assumed equal to SARS-CoV-2 (Wuhan) |
| | Rs4084 | 0.048 | Assumed equal to SARS-CoV-2 (Wuhan) |

| | | | | |
|---|---|---|---|---|
| | GX/P1E | 0.048 | | Assumed equal to SARS-CoV-2 (Wuhan) |
| | RaTG13 | 0.048 | | Assumed equal to SARS-CoV-2 (Wuhan) |
| *Transmission Rate* | SARS-CoV-2 (Wuhan) (by age) | 0-5:<br>5-10:<br>10-15:<br>15-20:<br>20-25:<br>25-30:<br>30-35:<br>35-40:<br>40-45:<br>45-50:<br>50-55:<br>55-60:<br>60-65:<br>65-70:<br>70-75:<br>75+: | 0.07506279<br>0.07506635<br>0.07507211<br>0.07508187<br>0.07509726<br>0.07512216<br>0.07516265<br>0.07522881<br>0.07533744<br>0.07551206<br>0.07578919<br>0.07622652<br>0.07691883<br>0.07800548<br>0.07954659<br>0.0843165 | Calculated from other parameters. |
| | SARS-CoV-2 (Alpha) (by age) | 0-5:<br>5-10:<br>10-15:<br>15-20:<br>20-25:<br>25-30:<br>30-35:<br>35-40:<br>40-45:<br>45-50:<br>50-55:<br>55-60:<br>60-65:<br>65-70:<br>70-75:<br>75+: | 0.13077016<br>0.13077264<br>0.13077666<br>0.13078346<br>0.13079419<br>0.13081155<br>0.13083977<br>0.1308859<br>0.13096163<br>0.13108337<br>0.13127656<br>0.13158145<br>0.1320641<br>0.13282165<br>0.13389604<br>0.13722138 | Calculated from other parameters. |
| | SARS-CoV-2 (Delta) (by age) | 0-5:<br>5-10:<br>10-15:<br>15-20:<br>20-25:<br>25-30:<br>30-35:<br>35-40:<br>40-45:<br>45-50:<br>50-55:<br>55-60:<br>60-65:<br>65-70:<br>70-75:<br>75+: | 0.25953183<br>0.25953567<br>0.25954188<br>0.25955239<br>0.25956897<br>0.25959581<br>0.25963943<br>0.25971072<br>0.25982776<br>0.2600159<br>0.26031449<br>0.2607857<br>0.26153164<br>0.26270244<br>0.26436293<br>0.26950229 | Calculated from other parameters. |
| | SARS-CoV-2 (Omicron) (by age) | 0-5:<br>5-10:<br>10-15:<br>15-20:<br>20-25:<br>25-30:<br>30-35:<br>35-40:<br>40-45:<br>45-50:<br>50-55:<br>55-60:<br>60-65:<br>65-70: | 0.69101496<br>0.69102064<br>0.69102982<br>0.69104538<br>0.69106991<br>0.69110961<br>0.69117416<br>0.69127964<br>0.69145282<br>0.69173121<br>0.69217301<br>0.69287023<br>0.69397394<br>0.6957063 | Calculated from other parameters. |

| | | | |
|---|---|---|---|
| | 70-75: | 0.69816321 | |
| | 75+: | 0.70576758 | |
| SARS-CoV-1 (by age) | 0-5: | 0.07506279 | Assumed equal to SARS-CoV-2 (Wuhan) |
| | 5-10: | 0.07506635 | |
| | 10-15: | 0.07507211 | |
| | 15-20: | 0.07508187 | |
| | 20-25: | 0.07509726 | |
| | 25-30: | 0.07512216 | |
| | 30-35: | 0.07516265 | |
| | 35-40: | 0.07522881 | |
| | 40-45: | 0.07533744 | |
| | 45-50: | 0.07551206 | |
| | 50-55: | 0.07578919 | |
| | 55-60: | 0.07622652 | |
| | 60-65: | 0.07691883 | |
| | 65-70: | 0.07800548 | |
| | 70-75: | 0.07954659 | |
| | 75+: | 0.0843165 | |
| Rs4084 (by age) | 0-5: | 0.07506279 | Assumed equal to SARS-CoV-2 (Wuhan) |
| | 5-10: | 0.07506635 | |
| | 10-15: | 0.07507211 | |
| | 15-20: | 0.07508187 | |
| | 20-25: | 0.07509726 | |
| | 25-30: | 0.07512216 | |
| | 30-35: | 0.07516265 | |
| | 35-40: | 0.07522881 | |
| | 40-45: | 0.07533744 | |
| | 45-50: | 0.07551206 | |
| | 50-55: | 0.07578919 | |
| | 55-60: | 0.07622652 | |
| | 60-65: | 0.07691883 | |
| | 65-70: | 0.07800548 | |
| | 70-75: | 0.07954659 | |
| | 75+: | 0.0843165 | |
| GX/P1E (by age) | 0-5: | 0.07506279 | Assumed equal to SARS-CoV-2 (Wuhan) |
| | 5-10: | 0.07506635 | |
| | 10-15: | 0.07507211 | |
| | 15-20: | 0.07508187 | |
| | 20-25: | 0.07509726 | |
| | 25-30: | 0.07512216 | |
| | 30-35: | 0.07516265 | |
| | 35-40: | 0.07522881 | |
| | 40-45: | 0.07533744 | |
| | 45-50: | 0.07551206 | |
| | 50-55: | 0.07578919 | |
| | 55-60: | 0.07622652 | |
| | 60-65: | 0.07691883 | |
| | 65-70: | 0.07800548 | |
| | 70-75: | 0.07954659 | |
| | 75+: | 0.0843165 | |
| RaTG13 (by age) | 0-5: | 0.07506279 | Assumed equal to SARS-CoV-2 (Wuhan) |
| | 5-10: | 0.07506635 | |
| | 10-15: | 0.07507211 | |
| | 15-20: | 0.07508187 | |
| | 20-25: | 0.07509726 | |
| | 25-30: | 0.07512216 | |
| | 30-35: | 0.07516265 | |
| | 35-40: | 0.07522881 | |
| | 40-45: | 0.07533744 | |
| | 45-50: | 0.07551206 | |
| | 50-55: | 0.07578919 | |
| | 55-60: | 0.07622652 | |
| | 60-65: | 0.07691883 | |
| | 65-70: | 0.07800548 | |
| | 70-75: | 0.07954659 | |
| | 75+: | 0.0843165 | |

| | | | | |
|---|---|---|---|---|
| *Infection Mortality Rate* | SARS-CoV-2 (Wuhan) (by age) | 0-5:<br>5-10:<br>10-15:<br>15-20:<br>20-25:<br>25-30:<br>30-35:<br>35-40:<br>40-45:<br>45-50:<br>50-55:<br>55-60:<br>60-65:<br>65-70:<br>70-75:<br>75+: | 0.00000364<br>0.0000059<br>0.00000956<br>0.00001575<br>0.00002552<br>0.00004133<br>0.00006703<br>0.00010903<br>0.000178<br>0.00028885<br>0.00046478<br>0.00074241<br>0.00118192<br>0.00187175<br>0.00285011<br>0.0058782 | [12] |
| | SARS-CoV-2 (Alpha) (by age) | 0-5:<br>5-10:<br>10-15:<br>15-20:<br>20-25:<br>25-30:<br>30-35:<br>35-40:<br>40-45:<br>45-50:<br>50-55:<br>55-60:<br>60-65:<br>65-70:<br>70-75:<br>75+: | 0.00000364<br>0.0000059<br>0.00000956<br>0.00001575<br>0.00002552<br>0.00004133<br>0.00006703<br>0.00010903<br>0.000178<br>0.00028885<br>0.00046478<br>0.00074241<br>0.00118192<br>0.00187175<br>0.00285011<br>0.0058782 | [12] |
| | SARS-CoV-2 (Delta) (by age) | 0-5:<br>5-10:<br>10-15:<br>15-20:<br>20-25:<br>25-30:<br>30-35:<br>35-40:<br>40-45:<br>45-50:<br>50-55:<br>55-60:<br>60-65:<br>65-70:<br>70-75:<br>75+: | 0.00000364<br>0.0000059<br>0.00000956<br>0.00001575<br>0.00002552<br>0.00004133<br>0.00006703<br>0.00010903<br>0.000178<br>0.00028885<br>0.00046478<br>0.00074241<br>0.00118192<br>0.00187175<br>0.00285011<br>0.0058782 | [12] |
| | SARS-CoV-2 (Omicron) (by age) | 0-5:<br>5-10:<br>10-15:<br>15-20:<br>20-25:<br>25-30:<br>30-35:<br>35-40:<br>40-45:<br>45-50:<br>50-55:<br>55-60:<br>60-65:<br>65-70:<br>70-75:<br>75+: | 0.00000364<br>0.0000059<br>0.00000956<br>0.00001575<br>0.00002552<br>0.00004133<br>0.00006703<br>0.00010903<br>0.000178<br>0.00028885<br>0.00046478<br>0.00074241<br>0.00118192<br>0.00187175<br>0.00285011<br>0.0058782 | [12] |
| | SARS-CoV-1 (by age) | 0-5:<br>5-10: | 0.00006223<br>0.00010077 | Inferred from non-linear least squares model (Supplementary |

| | | | | |
|---|---|---|---|---|
| | | 10-15: | 0.00016279 | Figure 12) with population estimate taken from [13] |
| | | 15-20: | 0.00026709 | |
| | | 20-25: | 0.00042973 | |
| | | 25-30: | 0.00068816 | |
| | | 30-35: | 0.00109603 | |
| | | 35-40: | 0.00173207 | |
| | | 40-45: | 0.00270132 | |
| | | 45-50: | 0.00409076 | |
| | | 50-55: | 0.00595211 | |
| | | 55-60: | 0.0082628 | |
| | | 60-65: | 0.01090161 | |
| | | 65-70: | 0.01361388 | |
| | | 70-75: | 0.0159554 | |
| | | 75+: | 0.01908244 | |
| | Rs4084 (by age) | 0-5: | 0.00006223 | Assumed equal to SARS-CoV-1 |
| | | 5-10: | 0.00010077 | |
| | | 10-15: | 0.00016279 | |
| | | 15-20: | 0.00026709 | |
| | | 20-25: | 0.00042973 | |
| | | 25-30: | 0.00068816 | |
| | | 30-35: | 0.00109603 | |
| | | 35-40: | 0.00173207 | |
| | | 40-45: | 0.00270132 | |
| | | 45-50: | 0.00409076 | |
| | | 50-55: | 0.00595211 | |
| | | 55-60: | 0.0082628 | |
| | | 60-65: | 0.01090161 | |
| | | 65-70: | 0.01361388 | |
| | | 70-75: | 0.0159554 | |
| | | 75+: | 0.01908244 | |
| | GX/P1E (by age) | 0-5: | 0.00000364 | Assumed equal to SARS-CoV-2 (Wuhan) |
| | | 5-10: | 0.0000059 | |
| | | 10-15: | 0.00000956 | |
| | | 15-20: | 0.00001575 | |
| | | 20-25: | 0.00002552 | |
| | | 25-30: | 0.00004133 | |
| | | 30-35: | 0.00006703 | |
| | | 35-40: | 0.00010903 | |
| | | 40-45: | 0.000178 | |
| | | 45-50: | 0.00028885 | |
| | | 50-55: | 0.00046478 | |
| | | 55-60: | 0.00074241 | |
| | | 60-65: | 0.00118192 | |
| | | 65-70: | 0.00187175 | |
| | | 70-75: | 0.00285011 | |
| | | 75+: | 0.0058782 | |
| | RaTG13 (by age) | 0-5: | 0.00000364 | Assumed equal to SARS-CoV-2 (Wuhan) |
| | | 5-10: | 0.0000059 | |
| | | 10-15: | 0.00000956 | |
| | | 15-20: | 0.00001575 | |
| | | 20-25: | 0.00002552 | |
| | | 25-30: | 0.00004133 | |
| | | 30-35: | 0.00006703 | |
| | | 35-40: | 0.00010903 | |
| | | 40-45: | 0.000178 | |
| | | 45-50: | 0.00028885 | |
| | | 50-55: | 0.00046478 | |
| | | 55-60: | 0.00074241 | |
| | | 60-65: | 0.00118192 | |
| | | 65-70: | 0.00187175 | |
| | | 70-75: | 0.00285011 | |
| | | 75+: | 0.0058782 | |
| *Immunity Waning Rate* | SARS-CoV-2 (Wuhan) | 0.003954 | | Inferred through epidemiological model fitted to data in [11] |
| | SARS-CoV-2 (Alpha) | 0.003627 | | Inferred through epidemiological model fitted to data in [11] |

| | | |
|---|---|---|
| SARS-CoV-2 (Delta) | 0.007329 | Inferred through epidemiological model fitted to data in [11] |
| SARS-CoV-2 (Omicron) | 0.01801 | Inferred through epidemiological model fitted to data in [11] |
| SARS-CoV-1 | 0.003954 | Assumed equal to SARS-CoV-2 (Wuhan) |
| Rs4084 | 0.003954 | Assumed equal to SARS-CoV-2 (Wuhan) |
| GX/P1E | 0.003954 | Assumed equal to SARS-CoV-2 (Wuhan) |
| RaTG13 | 0.003954 | Assumed equal to SARS-CoV-2 (Wuhan) |
